# Evaluating ChatGPT’s Performance in Responding to Questions About Endoscopic Procedures for Patients

**DOI:** 10.1101/2023.05.31.23290800

**Authors:** Hassam Ali, Pratik Patel, Itegbemie Obaitan, Babu P. Mohan, Amir Humza Sohail, Lucia Smith-Martinez, Karrisa Lambert, Manesh Kumar Gangwani, Jeffrey J. Easler, Douglas G. Adler

## Abstract

**Background and aims:** We aimed to assess the accuracy, completeness, and consistency of ChatGPT’s responses to frequently asked questions concerning the management and care of patients receiving endoscopic procedures and to compare its performance to Generative Pre-trained Transformer 4 (GPT-4) in providing emotional support.

**Methods:** Frequently asked questions (N = 117) about esophagogastroduodenoscopy (EGD), colonoscopy, endoscopic ultrasound (EUS), and endoscopic retrograde cholangiopancreatography (ERCP) were collected from professional societies, institutions, and social media. ChatGPT’s responses were generated and graded by board-certified gastroenterologists and advanced endoscopists. Emotional support questions were assessed by a psychiatrist.

**Results:** ChatGPT demonstrated high accuracy in answering questions about EGD (94.8% comprehensive or correct but insufficient), colonoscopy (100% comprehensive or correct but insufficient), ERCP (91% comprehensive or correct but insufficient), and EUS (87% comprehensive or correct but insufficient). No answers were deemed entirely incorrect (0%). Reproducibility was significant across all categories. ChatGPT’s emotional support performance was inferior to the newer GPT-4 model.

**Conclusion:** ChatGPT provides accurate and consistent responses to patient questions about common endoscopic procedures and demonstrates potential as a supplementary information resource for patients and healthcare providers.

## Introduction

Endoscopy remains vital in managing gastrointestinal diseases; it provides essential diagnostic and therapeutic interventions for innumerable gastrointestinal conditions. [1] In 2020, approximately 20 million endoscopic procedures were performed in the United States, highlighting the extensive reliance on these procedures in clinical settings. [2]

The most common endoscopic procedures include esophagogastroduodenoscopy (EGD), colonoscopy, endoscopic ultrasound (EUS), and endoscopic retrograde cholangiopancreatography (ERCP). Due to the widespread nature of gastrointestinal disorders and the significance of endoscopic procedures in addressing these issues, patients will frequently have questions regarding these techniques.

Artificial intelligence (AI) has made remarkable strides in natural language processing (NLP) in recent years. [3] Models such as ChatGPT (Generative Pre-trained Transformer) and GPT-4 (Generative Pre-trained Transformer-4), developed by OpenAI, an artificial intelligence research organization based in San Francisco, California, have demonstrated potential for various healthcare applications. [4] These models have been utilized in tasks including responding to medical student examination queries, creating basic medical reports, and offering information on various health-related subjects. [5, 6, 7] ChatGPT could potentially serve as a supplementary information resource for patients, improving patient education and outcomes. [8] Nevertheless, concerns persist about ChatGPT’s ability to provide accurate and comprehensive responses to detailed medical questions.

No current literature specifically investigates ChatGPT’s capabilities in addressing questions concerning common endoscopic procedures. Our study aims to assess the precision, comprehensiveness, and reliability of ChatGPT’s answers to common queries about patient care and management regarding endoscopic procedures. Moreover, we will compare the performance of ChatGPT (freely accessible) to GPT-4 (paid subscription/limited access) when responding to questions posed by patients, as this comparison could reveal further insight into its potential role as a virtual assistant for patients and healthcare providers in the realm of endoscopic procedures.

## Methods

### Data source

FAQs on endoscopic procedures were sourced from professional societies and institutional websites (Supplemental file 1). Excluding repetitive or unclear questions, we curated 117 on endoscopic procedures (EGD, colonoscopy, EUS, ERCP; Supplemental Table 1-4). Additionally, we tested ChatGPT and GPT-4’s psychological support capability with 16 questions (Supplemental Table 5), evaluated by a certified psychiatrist (LSM).

### Response generation

ChatGPT, an enhanced GPT-3.5 model introduced in November 2022, incorporates user feedback and restrictions for safer and relevant responses. Two authors input questions twice separately into ChatGPT’s March 23 version for reproducibility [8]. GPT-4, a subscription-based model, was not utilized for primary analysis [4].

### Grading of questions

Responses for EGD/Colonoscopy were graded by certified gastroenterologists (BPM and PP), while those for EUS/ERCP by advanced endoscopists (IO and JE). A grading system assessed response accuracy and comprehensiveness, with a third reviewer (KL) resolving discrepancies [8].

### Emotional support questions and responses

ChatGPT’s performance on emotional support was examined using modified FAQs (Supplemental Table 5). The responses were graded by a certified psychiatrist (LSM), ranging from “Not Comprehensive” to “Extremely Comprehensive”.

### Statistical analysis

Reproducibility, measured by the uniformity of two similarly-graded responses, was evaluated. Disparate responses were graded, with those falling into separate grading groups deemed significantly different. ChatGPT’s performance on emotional support questions was compared to GPT-4. Grading proportions for each endoscopic procedure domain were calculated as percentages (N%). Analysis used STATA (version 16.1).

The methodology has been described in detail in Supplementary file 1.

## Results

ChatGPT displayed high levels of precision when answering questions about EGD (N =39), colonoscopy (N =22), ERCP (N = 28), and EUS (N = 28) about treatment, lifestyle/aftercare, basic knowledge, and others (Figure 1).

**Figure 1:**
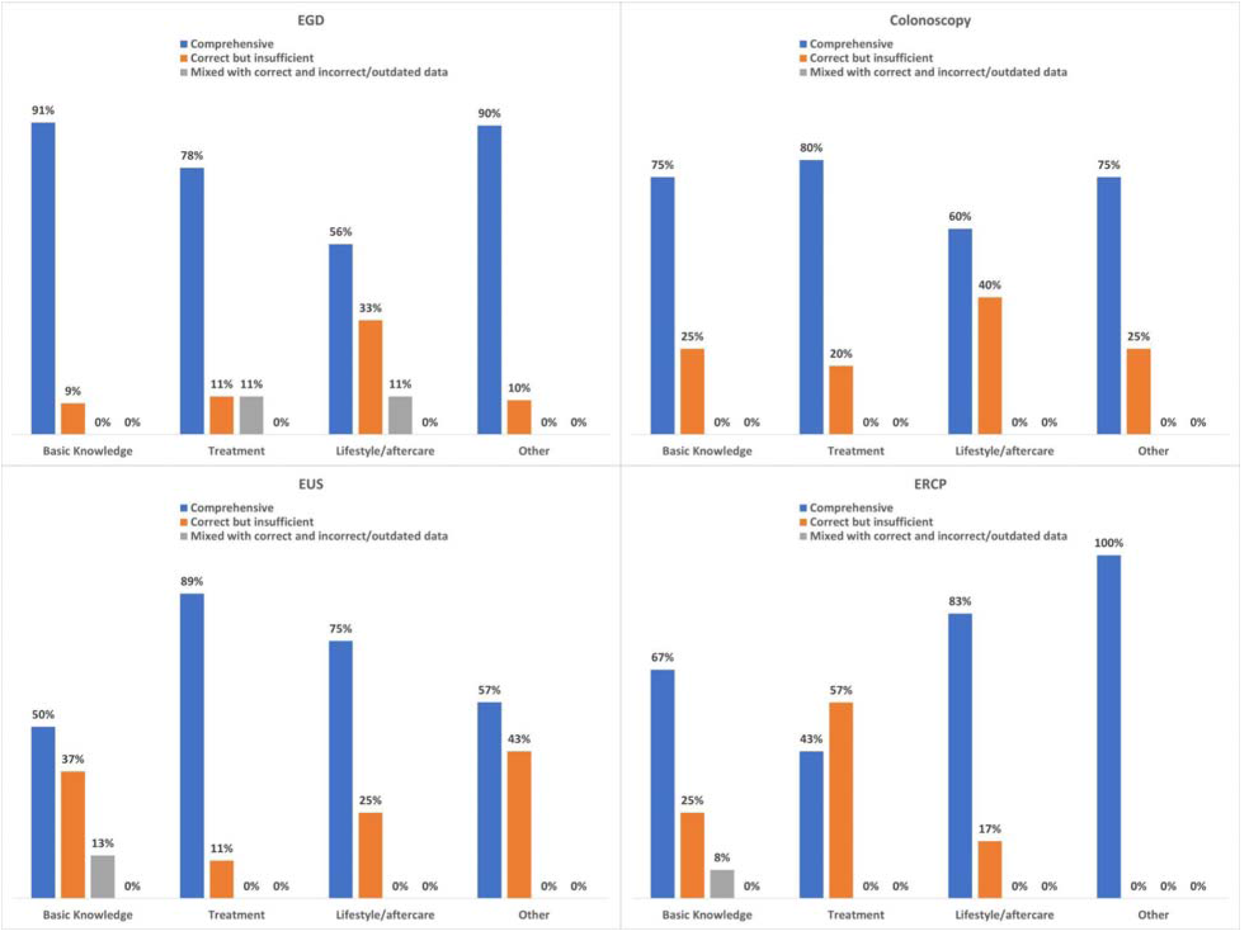
Grade of responses by the ChatGPT language model to questions related to endoscopic procedures.

**Figure 2:**
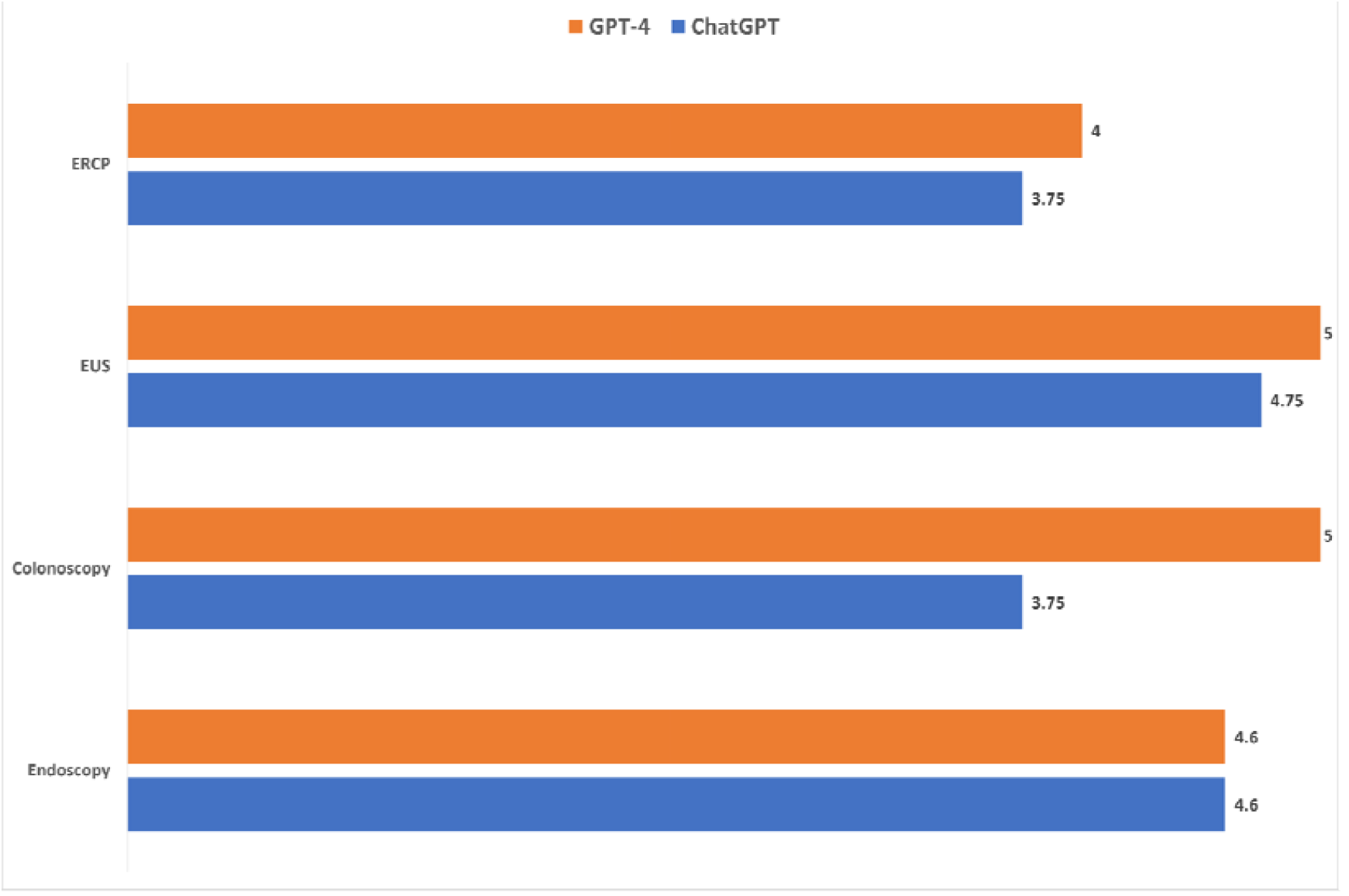
Responses to emotional support statements by ChatGPT and GPT-4 language models Not Comprehensive: The answer provided minimal information or needed to address the question adequately. Somewhat Comprehensive: The answer provided some information but left out important details. Moderately Comprehensive: The answer covered most aspects of the question but may have lacked depth or specificity in certain areas. Very Comprehensive: The answer addressed all aspects of the question and provided detailed information. Extremely Comprehensive: The answer was exhaustive, covering all aspects of the question with great detail and specificity.

### Frequently asked questions about EGD

The percentage of answers considered comprehensive or correct but insufficient was 94.8% or above for the “basic knowledge,” “treatment,” “lifestyle/aftercare,” and “others” categories (Supplementary Table 1). No answers from ChatGPT were deemed entirely incorrect.

Reproducibility was significant across all categories, with 100% (39/39) of all questions generating two comparable answers. The Reproducibility within specific categories is displayed in Table 1.

**Table 1.**
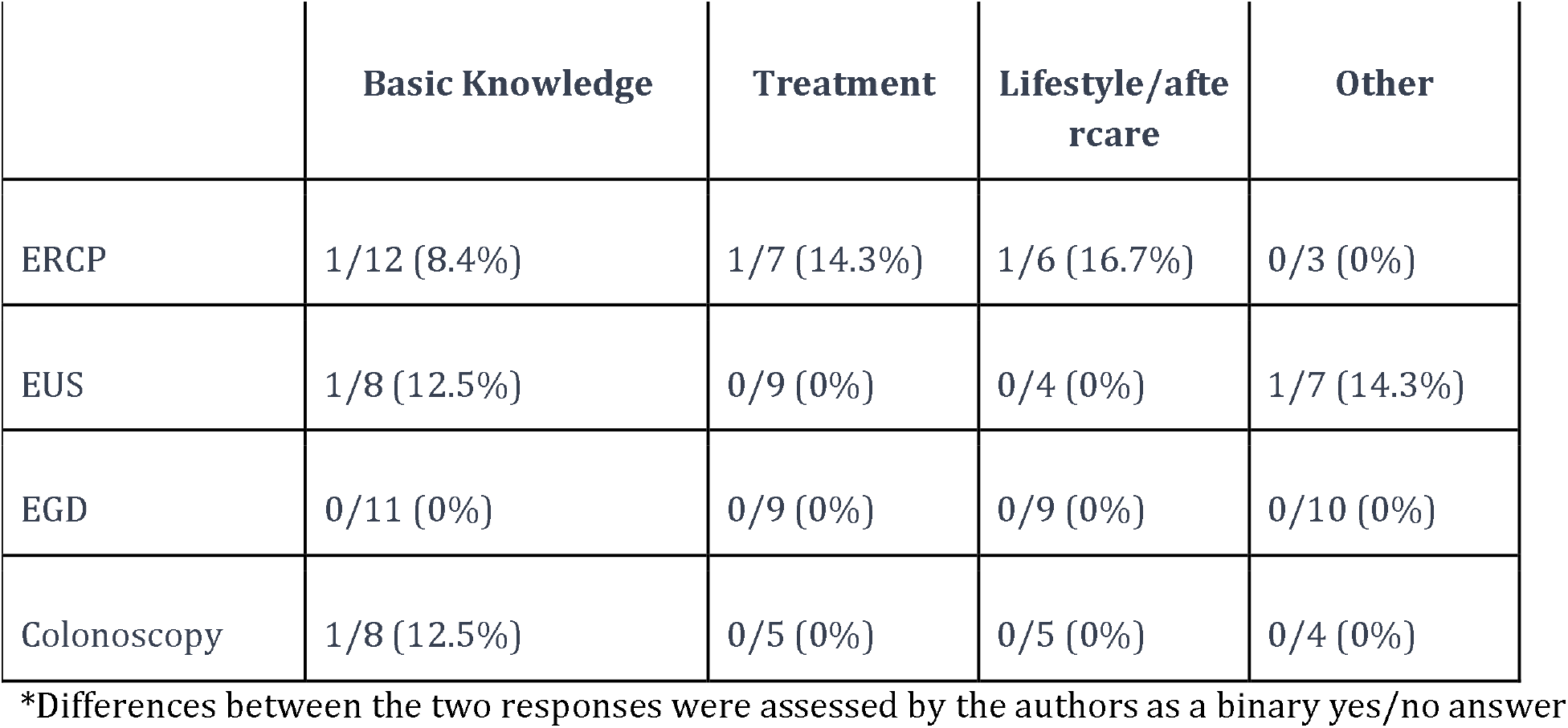
Percentage of questions with significantly different between the two responses.

|  | **Basic Knowledge** | **Treatment** | **Lifestyle/aftercare** | **Other** |
| --- | --- | --- | --- | --- |
| ERCP | 1/12 (8.4%) | 1/7 (14.3%) | 1/6 (16.7%) | 0/3 (0%) |
| EUS | 1/8 (12.5%) | 0/9 (0%) | 0/4 (0%) | 1/7 (14.3%) |
| EGD | 0/11 (0%) | 0/9 (0%) | 0/9 (0%) | 0/10 (0%) |
| Colonoscopy | 1/8 (12.5%) | 0/5 (0%) | 0/5 (0%) | 0/4 (0%) |

\*Differences between the two responses were assessed by the authors as a binary yes/no answer

### Frequently asked questions about Colonoscopy

The percentage of answers considered comprehensive or correct but insufficient was 100 % or above for the “basic knowledge,” “treatment,” “lifestyle/aftercare,” and “others” categories (Supplementary Table 2). No answers from ChatGPT were deemed entirely incorrect. Reproducibility was significant across all categories, with 95.4% (21/22) of all questions generating two comparable answers. The Reproducibility within specific categories is displayed in Table 1.

### Frequently asked questions about ERCP

The percentage of answers considered comprehensive or correct but insufficient was 91% or above for the “basic knowledge,” “treatment,” “lifestyle/aftercare,” and “others” categories (Supplementary Table 3). No answers from ChatGPT were deemed entirely incorrect. Reproducibility was significant across all categories, with 89.2% (25/28) of all questions generating two comparable answers. The Reproducibility within specific categories is displayed in Table 1.

### Frequently asked questions about EUS

The percentage of answers considered comprehensive or correct but insufficient was 87% or above for the “basic knowledge,” “treatment,” “lifestyle/aftercare,” and “others” categories (Supplementary Table 4). No answers from ChatGPT were deemed entirely incorrect. Reproducibility was significant across all categories, with 92.8% (26/28) of all questions generating two comparable answers. The reproducibility within specific categories is displayed in Table 1.

### Emotional support questions about endoscopic procedures

The responses to emotional support questions were graded from 1-5 based on the level of comprehensiveness (Figure 1). Both LLMs (ChatGPT and GPT-4) performed adequately, with all responses being moderate to extremely comprehensive. GPT-4 outperformed ChatGPT responses to emotional questions (Supplementary Table 5). No answers from either LLM were deemed noncomprehensive.

## Discussion

This study assessed the precision and consistency of ChatGPT in addressing patient inquiries about endoscopic procedures. ChatGPT generated accurate and relevant responses to these procedures and provided comprehensive information to patients, performing better with basic procedures like EGD and colonoscopy compared to EUS/ERCP. ChatGPT also effectively addressed emotional concerns, showcasing empathy and understanding [15].

Global differences in endoscopic guidelines weren’t evaluated. GPT-4 was not examined due to accessibility constraints. ChatGPT could be a supplementary patient resource, enhancing gastroenterological procedure comprehension.

Health literacy is vital for patients undergoing GI endoscopic procedures [9,10]. Despite the need for accessible and accurate information, obtaining easy-to-understand resources can be a struggle. ChatGPT can address this issue by delivering health information conversationally, simplifying complex medical jargon [11, 12], potentially leading to better patient understanding [13]. ChatGPT can support healthcare providers by generating responses to routine patient inquiries, potentially saving time for more complex cases. The accuracy of the responses varies, and with technological improvements, this could increase, possibly boosting provider productivity [14]. ChatGPT and GPT-4 showed empathetic responses to emotional questions. Further research is needed to evaluate this capability, including comprehension of complex inquiries and cultural adaptation.

This study’s main strengths include the comprehensive collection of inquiries from authoritative sources. However, ChatGPT has limitations. A few questions got comprehensive responses, hinting at its role as a supplemental tool rather than a replacement for healthcare providers. Discrepancies (<25%) among reviewers demonstrate variation in expert opinions. ChatGPT’s training data, limited to 2021, may lead to outdated responses. Its training data’s quality remains under review, affecting reliability. Furthermore, ChatGPT struggled with specifics like lab cut-offs or treatment durations. Reviewers’ awareness of ChatGPT might have led to stricter grading, potentially underestimating its performance. Finally, globally varying guidelines could lead to confusion or harm if not correctly specified. Further refinement is needed to enhance data reliability and specificity.

ChatGPT can augment healthcare providers, assisting patients with pertinent questions. Our study examined the accuracy and reproducibility of ChatGPT’s responses to common patient inquiries on GI endoscopic procedures. ChatGPT frequently provided accurate, albeit sometimes incomplete, responses. The model’s advice, varying across regions, should not be solely trusted.

## Data Availability

All data produced in the present work are contained in the manuscript

