## Supplemental Tables for "Evaluating ChatGPT’s Performance in Responding to Questions About Endoscopic Procedures for Patients"

Supplemental Table 1. Grading of the responses from the ChatGPT Language Model for Esophagogastroduodenoscopy (EGD)-related questions……….2

Supplemental Table 2. Grading of the responses from the ChatGPT Language Model for colonoscopy-related questions………………………………….34

Supplemental Table 3. Grading of the responses from the ChatGPT Language Model for endoscopic ultrasound (EUS)-related questions………………..53

Supplemental Table 4. Grading of the responses from the ChatGPT Language Model for Endoscopic retrograde cholangiopancreatography (ERCP)-related questions……………………………………………………………………………………………………………………………………………………….85

Supplemental Table 5: Grading responses to emotional support questions by ChatGPT and GPT-4 models………………………………………………..112

Sources for questions …………………………………………………………………………………………………………………………………………132

Methods in detail ……………………………………………………………………………………………………………………………………………..133

Supplemental Table 1. Grading of the responses from the ChatGPT Language Model for Esophagogastroduodenoscopy (EGD)-related questions

| **Basic Knowledge:** | 1st response | 2nd response | Reviewer 1 grades 1. Comprehensive 2. Correct but inadequate 3. Mixed with correct and incorrect/outdated data 4. Completely incorrect | Reviewer 1 grades 1. Comprehensive 2. Correct but inadequate 3. Mixed with correct and incorrect/outdated data 4. Completely incorrect | Reviewer 3 grades for the responses with discrepancies |
| --- | --- | --- | --- | --- | --- |
| What is an esophagogastroduodenoscopy ? | An esophagogastroduodenoscopy is a medical procedure that involves the use of an instrument called an endoscope to examine the inside of a person's body. The endoscope is a flexible tube with a light and camera attached to the end, which allows a doctor to see inside the body without making a large incision. Endoscopies are commonly used to examine the digestive system, including the esophagus, stomach, and intestines. The procedure can be used to diagnose a variety of conditions, such as ulcers, inflammation, or cancer. It can also be used to perform certain treatments, such as removing polyps or taking tissue samples for biopsy. During the procedure, the patient is typically given sedation to help them relax and minimize discomfort. The endoscope is then inserted through the mouth or anus and guided through the digestive tract to the area being examined. The doctor can view the images captured by the camera on a monitor and may take samples or perform other procedures as needed. The procedure usually takes around 30 minutes to an hour, and the patient can typically go home the same day. | similar | 1 | 2 | Comprehensive |
| How is an esophagogastroduodenoscopy performed? | An esophagogastroduodenoscopy is a medical procedure in which a thin, flexible tube with a camera attached to it, called an endoscope, is inserted into the body to view the inside of organs or cavities. Here are the general steps involved in an esophagogastroduodenoscopy procedure: Preparation: Before the procedure, the patient may be given a sedative or anesthetic to help them relax or prevent discomfort. The patient may also be asked to fast for a certain period of time prior to the procedure. Insertion: The endoscope is inserted into the body through a natural opening, such as the mouth, anus, or nose, or through a small incision. The doctor guides the endoscope through the body to the area being examined. Examination: As the endoscope is moved through the body, the camera attached to it transmits images to a monitor, allowing the doctor to see inside the body. The doctor may also use tools attached to the endoscope to take tissue samples or perform other procedures. Removal: Once the examination is complete, the endoscope is slowly withdrawn from the body. Recovery: After the procedure, the patient may need to stay in a recovery area for a short period of time while the effects of any sedatives or anesthesia wear off. The doctor will usually discuss the results of the examination with the patient and recommend any necessary follow-up care. It's important to note that the specific details of an esophagogastroduodenoscopy procedure may vary depending on the type of esophagogastroduodenoscopy being performed and the reason for the examination. The doctor will provide specific instructions and information to the patient prior to the procedure. | similar | 1 | 1 | Comprehensive |
| What types of endoscopies are there? | esophagogastroduodenoscopy is a medical procedure that involves using an endoscope, a long, flexible tube with a light and camera attached to it, to examine the inside of the body. There are many different types of endoscopies, each with a specific purpose and location in the body. Here are some examples: Upper esophagogastroduodenoscopy (esophagogastroduodenoscopy or EGD): This type of esophagogastroduodenoscopy is used to examine the upper gastrointestinal tract, including the esophagus, stomach, and the first part of the small intestine (duodenum). Colonoscopy: This is an endoscopic examination of the large intestine (colon) and rectum. It is commonly used to screen for colon cancer and other colon-related diseases. Bronchoscopy: This procedure involves examining the airways and lungs using an endoscope inserted through the nose or mouth. Cystoscopy: This endoscopic examination is used to visualize the bladder and urethra, and is commonly used to diagnose urinary tract infections and other urological disorders. Hysteroscopy: This is an endoscopic examination of the uterus, typically performed to diagnose and treat abnormal uterine bleeding, infertility, or other conditions affecting the reproductive system. Enteroscopy: This type of esophagogastroduodenoscopy examines the small intestine, using a long, thin endoscope inserted through the mouth or anus. These are just a few examples of the many different types of endoscopies available, and the specific type of esophagogastroduodenoscopy used will depend on the part of the body being examined and the reason for the examination. | similar | 3 | 2 | Correct but inadequate |
| What are the risks associated with esophagogastroduodenoscopy ? | esophagogastroduodenoscopy is a medical procedure that uses an instrument called an endoscope to examine the interior of the body, typically the digestive tract, respiratory tract, or urinary tract. Like any medical procedure, esophagogastroduodenoscopy carries some risks, although these risks are generally low. Here are some potential risks associated with esophagogastroduodenoscopy : Bleeding: esophagogastroduodenoscopy can cause bleeding if the scope tears the lining of the digestive tract or another organ. This risk is higher in patients who have underlying medical conditions that affect blood clotting. Infection: esophagogastroduodenoscopy can introduce bacteria or other microorganisms into the body, which can lead to infection. This risk is higher if the endoscope is not properly cleaned and disinfected between procedures. Perforation: esophagogastroduodenoscopy can cause a tear or hole in the digestive tract or other organ. This is a rare but serious complication that can lead to infection or other complications. Adverse reaction to anesthesia: If the patient receives sedation or anesthesia during the procedure, there is a risk of an allergic reaction or other adverse reaction to the medication. Other risks: Other potential risks associated with esophagogastroduodenoscopy include breathing difficulties, heart problems, and reactions to the contrast material used during certain types of endoscopic procedures. It is important to discuss the risks and benefits of esophagogastroduodenoscopy with your doctor before undergoing the procedure. Your doctor can help you determine if esophagogastroduodenoscopy is the best option for your individual needs and can help you understand the potential risks involved. | similar | 1 | 1 | Comprehensive |
| Do I need to fast before an esophagogastroduodenoscopy ? | Yes, it is generally recommended to fast before an esophagogastroduodenoscopy . This is because during an esophagogastroduodenoscopy , a doctor will insert a flexible tube with a camera on the end into your digestive system to examine your esophagus, stomach, and small intestine. Fasting helps to ensure that your stomach and small intestine are empty, which can improve the clarity of the images obtained during the esophagogastroduodenoscopy and reduce the risk of complications such as vomiting or aspiration. Your doctor will provide you with specific instructions on how long to fast before the procedure, which may vary depending on the type of esophagogastroduodenoscopy being performed and your individual medical history. In general, you may be asked to stop eating and drinking for 6 to 12 hours before the procedure. It is important to follow your doctor's instructions carefully to ensure a safe and successful esophagogastroduodenoscopy . | similar | 1 | 1 | Comprehensive |
| How long does an esophagogastroduodenoscopy take? | The length of an esophagogastroduodenoscopy procedure can vary depending on the type of esophagogastroduodenoscopy being performed and the individual circumstances of the patient. Generally, an esophagogastroduodenoscopy procedure takes between 15 minutes to an hour to complete. However, the time can vary based on the complexity of the procedure and the patient's health condition. For example, a diagnostic upper esophagogastroduodenoscopy , which is used to examine the esophagus, stomach, and upper small intestine, typically takes around 15 to 30 minutes to complete. A colonoscopy, which is used to examine the large intestine and rectum, can take between 30 minutes to an hour to complete. If polyps or other abnormalities are found during the procedure, additional time may be required for their removal or biopsy. It is important to note that patients undergoing an esophagogastroduodenoscopy will need to be sedated or given anesthesia, which can prolong the recovery time after the procedure. Additionally, patients may need to remain under observation for a period of time following the procedure to ensure there are no complications. | similar | 1 | 2 | Comprehensive |
| Will I be sedated during the esophagogastroduodenoscopy ? | Whether or not you will be sedated during an esophagogastroduodenoscopy depends on the specific procedure you are having and the preferences of your healthcare provider. In some cases, a mild sedative may be given to help you relax and make you more comfortable during the procedure. In other cases, a deeper sedation or anesthesia may be necessary, especially if the procedure is more invasive or if you have a history of anxiety or discomfort during medical procedures. It's important to discuss your concerns and preferences with your healthcare provider before the procedure so that they can make a recommendation for the most appropriate sedation or anesthesia option for you. Additionally, they will provide you with instructions on how to prepare for the procedure and any necessary restrictions on eating or drinking before the procedure. | similar | 1 | 1 | Comprehensive |
| How is the endoscope inserted into the body? | An endoscope is a flexible or rigid instrument used to examine the inside of the body. The method of insertion depends on the type of endoscope being used and the area of the body being examined. Typically, the procedure starts with the patient being given anesthesia or a sedative to help them relax. Then, the endoscope is inserted through a natural opening in the body, such as the mouth, nose, or anus. In some cases, a small incision may be made to allow the endoscope to be inserted. The endoscope is guided through the body by a trained medical professional, who views the images transmitted by the camera on the endoscope on a monitor. The doctor may use specialized tools on the endoscope to take biopsies, remove tissue or foreign objects, or perform other procedures. After the procedure, the endoscope is removed and the patient is monitored for a short time to ensure that there are no complications. | similar | 1 | 1 | Comprehensive |
| How long does it take for the sedative to wear off after the esophagogastroduodenoscopy ? | The length of time it takes for the sedative to wear off after an esophagogastroduodenoscopy can vary depending on the type and amount of sedative used, as well as individual factors such as age, weight, and overall health. However, in general, it can take several hours for the effects of the sedative to wear off completely. After the procedure, you will be taken to a recovery area where you will be monitored until the effects of the sedative have worn off sufficiently for you to be discharged. This typically takes around 1-2 hours, but may take longer in some cases. During the recovery period, it is important to follow your healthcare provider's instructions and avoid driving, operating machinery, or making important decisions until the sedative has completely worn off and you feel fully alert. You should also avoid eating or drinking until your healthcare provider has determined that it is safe to do so. | similar | 1 | 1 | Comprehensive |
| Can children have an esophagogastroduodenoscopy ? | Yes, children can have an esophagogastroduodenoscopy . An esophagogastroduodenoscopy is a medical procedure in which a doctor uses a thin, flexible tube with a camera and light at the end to look inside a person's body. Endoscopies can be used to examine the digestive tract, respiratory system, urinary system, or other parts of the body. Endoscopies can be performed on children, including infants, but the procedure may be modified to suit their age and size. For example, the doctor may use a smaller endoscope or use sedation to keep the child calm during the procedure. The exact approach will depend on the specific medical situation and the child's individual needs. It's important for parents to talk to their child's doctor about any concerns they may have regarding the esophagogastroduodenoscopy and to follow any preparation instructions provided. In general, an esophagogastroduodenoscopy is a safe and effective way to diagnose and treat certain medical conditions in children. | similar | 1 | 1 | Comprehensive |
| why do I need an esophagogastroduodenoscopy ? | An esophagogastroduodenoscopy is a medical procedure that allows doctors to examine the inside of your digestive tract using a thin, flexible tube with a camera and light source at the end. Endoscopies are commonly used to diagnose and treat a variety of gastrointestinal problems, including: Acid reflux or heartburn Difficulty swallowing or painful swallowing Abdominal pain Nausea and vomiting Unexplained weight loss Gastrointestinal bleeding Ulcers Polyps or tumors Inflammatory bowel disease Celiac disease Your doctor may recommend an esophagogastroduodenoscopy if you are experiencing any of these symptoms, or if you have a family history of gastrointestinal problems. It is important to follow your doctor's recommendations and undergo any necessary diagnostic tests to identify and treat any potential health issues. | similar | 1 | 2 | Comprehensive |
| **Treatment:** |  |  |  |  |  |
| Will I feel pain during the esophagogastroduodenoscopy ? | During an esophagogastroduodenoscopy , the doctor will use a long, thin, flexible tube with a camera and light at the end to examine your digestive tract. You will usually be given a sedative to help you relax and may also be given a local anesthetic to numb the area where the endoscope will be inserted. While most people do not feel any pain during the procedure, some people may experience discomfort or a mild burning sensation when the endoscope is passed through the throat or into the stomach. If you do experience pain, you should let the doctor know immediately so that they can adjust the position of the endoscope or give you additional medication to help manage your discomfort. It's important to remember that the discomfort or pain you may experience during an esophagogastroduodenoscopy is typically short-lived and generally outweighed by the benefits of having the procedure, which can help diagnose and treat a variety of digestive tract conditions. | similar | 1 | 2 | Comprehensive |
| How will the results of the esophagogastroduodenoscopy be used in my treatment? | The results of an esophagogastroduodenoscopy procedure will depend on the specific reason why the procedure was ordered. esophagogastroduodenoscopy is a diagnostic tool used by doctors to examine the inside of the body and diagnose any problems that may be present. Depending on the specific findings from the esophagogastroduodenoscopy , your doctor may recommend a number of different treatments or interventions. For example, if the esophagogastroduodenoscopy reveals that you have inflammation or damage in your digestive tract, your doctor may recommend medications to reduce inflammation, antibiotics to treat any infections, or changes to your diet to avoid foods that may be exacerbating the problem. If the esophagogastroduodenoscopy detects a growth or mass, your doctor may recommend a biopsy to determine if the growth is cancerous or not, and if it is, they may recommend surgery or other treatments to remove the cancer. Ultimately, the treatment plan that your doctor recommends will depend on the specific results of the esophagogastroduodenoscopy and the underlying condition that is being treated. It's important to discuss the findings with your doctor and ask any questions you may have about your treatment options. | similar | 1 | 1 | Comprehensive |
| How often will I need to have an esophagogastroduodenoscopy ? | The frequency at which you will need to have an esophagogastroduodenoscopy depends on several factors, including your medical history, current health status, and the reason for the procedure. esophagogastroduodenoscopy is a medical procedure that involves inserting a flexible tube with a camera and light source into your body to visualize and diagnose certain conditions. If you have a chronic condition that requires ongoing monitoring, such as gastroesophageal reflux disease (GERD) or Barrett's esophagus, you may need to have an esophagogastroduodenoscopy performed regularly, such as every one to three years. In some cases, more frequent monitoring may be required, depending on the severity of your condition and other factors. For diagnostic purposes, the frequency of esophagogastroduodenoscopy will depend on the specific medical condition being evaluated. For example, if you have a history of polyps in your colon or are at an increased risk for colon cancer, your doctor may recommend a colonoscopy every five to ten years. It's important to discuss your individual circumstances and medical history with your doctor to determine how often you need to have an esophagogastroduodenoscopy performed. They will be able to provide you with specific recommendations based on your unique needs. | similar | 1 | 3 | Mixed with correct and incorrect/outdated data |
| How do I prepare for an esophagogastroduodenoscopy ? | esophagogastroduodenoscopy is a medical procedure that involves examining the inside of your digestive system using an instrument called an endoscope. Here are some general guidelines on how to prepare for an esophagogastroduodenoscopy : Follow your doctor's instructions: Your doctor will give you specific instructions on how to prepare for your esophagogastroduodenoscopy . Follow these instructions carefully to ensure the procedure is successful and safe. Inform your doctor of any medications: Inform your doctor of any prescription or over-the-counter medications you are taking, including vitamins and supplements. You may need to stop taking some medications several days before the procedure. Avoid eating or drinking: You will be asked to avoid eating or drinking anything for several hours before the procedure, usually around six hours. Arrange for someone to accompany you: You may need someone to drive you home after the procedure, as you may still feel the effects of the sedative used during the esophagogastroduodenoscopy . Wear comfortable clothing: Wear loose, comfortable clothing that is easy to take off and put back on after the procedure. Follow the fasting guidelines: If you're undergoing an upper esophagogastroduodenoscopy , you will typically be instructed to fast for 6 to 8 hours before the procedure to ensure that the stomach is empty. If you're undergoing a colonoscopy, you will need to follow a clear liquid diet the day before and take laxatives to empty your bowel. Prepare for sedation: Depending on the type of esophagogastroduodenoscopy , you may receive sedation to help you relax or sleep during the procedure. Your doctor will inform you if you need to arrange for someone to drive you home afterward. Ask questions: Don't hesitate to ask your doctor any questions you have about the procedure or the preparation process. It's important to fully understand what to expect before and after the procedure. | similar | 1 | 1 | Comprehensive |
| What conditions can be diagnosed or treated with an esophagogastroduodenoscopy ? | An esophagogastroduodenoscopy (EGD), also known as an upper esophagogastroduodenoscopy , is a diagnostic and therapeutic procedure that allows a physician to examine the lining of the upper gastrointestinal tract, including the esophagus, stomach, and duodenum (the first part of the small intestine). The following conditions can be diagnosed or treated with an EGD: Gastroesophageal reflux disease (GERD) Hiatal hernia Ulcers (gastric or duodenal) Gastrointestinal bleeding Barrett's esophagus (a pre-cancerous condition) Esophageal cancer Gastric cancer Celiac disease Inflammatory bowel disease (Crohn's disease, ulcerative colitis) Strictures (narrowing of the esophagus, stomach or duodenum) Esophageal varices (dilated veins in the esophagus) Foreign bodies (objects that have been swallowed and are stuck in the upper GI tract) Polyps or tumors. During the procedure, the physician can take biopsies (small tissue samples) or remove polyps or other abnormal tissues for further analysis. EGD is a safe and effective procedure and can be performed on an outpatient basis under sedation. | similar | 1 | 2 | Comprehensive |
| What happens if the results of the esophagogastroduodenoscopy are inconclusive? | Esophagogastroduodenoscopy (EGD) is a diagnostic procedure used to examine the upper part of the digestive system, including the esophagus, stomach, and duodenum. In some cases, the results of an EGD may be inconclusive, meaning that the procedure did not provide a clear diagnosis. If the results of an EGD are inconclusive, the physician may recommend additional diagnostic tests or procedures to further investigate the cause of the symptoms. These tests may include imaging studies, such as an abdominal CT scan or MRI, or other specialized tests such as endoscopic ultrasound. In some cases, the physician may recommend a repeat EGD to obtain a more detailed view of the upper digestive tract. During a repeat EGD, the physician may take additional tissue samples for biopsy, which can provide more information about the underlying condition. It is important to follow the recommendations of your physician if the results of an EGD are inconclusive, as further testing may be necessary to obtain an accurate diagnosis and determine the best course of treatment. | similar | 1 | 1 | Comprehensive |
| How soon will I receive the results of the esophagogastroduodenoscopy ? | The timing of receiving the results of an esophagogastroduodenoscopy (EGD) can vary depending on several factors, including the policies and practices of the medical facility where the procedure was performed, the complexity of the findings, and the urgency of any necessary follow-up care. In general, you can expect to receive a preliminary report of the findings immediately after the procedure while you are still under the effects of sedation. This report may include initial impressions and any immediate interventions that were performed during the procedure. A more detailed and comprehensive report will typically be available within a few days after the procedure. This report will provide a more thorough analysis of the findings and any recommendations for further testing or treatment. It's best to consult with your doctor or healthcare provider to get a better idea of the specific timeline for receiving your EGD results based on your individual circumstances. | similar | 1 | 2 | Comprehensive |
| Are there any alternatives to having an esophagogastroduodenoscopy ? | Yes, there are alternative diagnostic tests that can be used instead of an esophagogastroduodenoscopy (EGD), also known as an upper endoscopy. Some possible alternatives include: Upper GI series: This is a diagnostic test that uses X-rays to visualize the esophagus, stomach, and small intestine. You will be asked to drink a contrast material (barium) that will coat the lining of your digestive tract and make it visible on the X-ray. While the test can identify some of the same conditions as an EGD, it may not be as accurate. Capsule endoscopy: This is a non-invasive procedure that uses a small, wireless camera pill that you swallow. As the capsule travels through your digestive tract, it takes pictures of the lining of your esophagus, stomach, and small intestine, which are then transmitted to a receiver worn on a belt or carried in a pocket. CT scan: A CT scan can be used to examine the esophagus, stomach, and small intestine. This diagnostic test uses X-rays and computer technology to create detailed images of the body. It may be useful in identifying masses, strictures or abnormalities in the esophagus, stomach or small intestine. Magnetic resonance imaging (MRI): This diagnostic test uses a strong magnetic field and radio waves to create detailed images of the body. MRI can also be used to evaluate the esophagus, stomach, and small intestine. It's important to note that the specific diagnostic test used will depend on the symptoms and condition being investigated. It is recommended to discuss your medical history and any concerns with your healthcare provider to determine the most appropriate diagnostic test. | similar | 2 | 2 | Correct but inadequate |
| How can I prepare for an esophagogastroduodenoscopy mentally and emotionally? | An esophagogastroduodenoscopy (EGD) is a medical procedure that involves the insertion of a thin, flexible tube with a camera at the end, called an endoscope, through your mouth and down your esophagus, stomach, and duodenum. It is used to diagnose and treat conditions such as acid reflux, ulcers, and gastrointestinal bleeding. To prepare yourself mentally and emotionally for an EGD, here are some steps you can take: Understand the procedure: It's important to know what to expect during the procedure. Ask your doctor or healthcare provider to explain the procedure in detail, including what will happen before, during, and after the EGD. This will help alleviate any anxiety or fears you may have. Talk to your healthcare provider: If you have any concerns or fears about the procedure, talk to your healthcare provider. They may be able to provide you with more information or recommend strategies to help you cope with your anxiety. Follow the instructions provided: Your healthcare provider will provide you with instructions on how to prepare for the EGD, which may include fasting for a certain amount of time before the procedure. Follow these instructions carefully to ensure that you are properly prepared for the procedure. Practice relaxation techniques: Relaxation techniques, such as deep breathing or meditation, can help calm your mind and body before the procedure. You may also want to consider listening to calming music or visualization techniques to help reduce any anxiety or stress. Bring a support person: Having a family member or friend accompany you to the procedure can provide emotional support and help you feel more comfortable. They can also drive you home after the procedure, as you will likely be sedated during the procedure. Remember, an EGD is a common and routine procedure that is performed thousands of times each day. By taking the time to prepare yourself mentally and emotionally, you can help ensure a smoother and more comfortable experience. | similar | 1 | 1 | Comprehensive |
| **Lifestyle/aftercare** |  |  |  |  |  |
| Can I eat and drink after an esophagogastroduodenoscopy ? | After an esophagogastroduodenoscopy (EGD), it is important to follow the instructions of your healthcare provider. In general, you may be advised to avoid eating or drinking for a period of time, usually a few hours, after the procedure. This is to allow your throat and digestive system to recover from the effects of the procedure and to avoid any potential complications, such as choking or aspiration. Your healthcare provider may also advise you to avoid certain foods or drinks for a period of time after the procedure, such as spicy or acidic foods, alcohol, or caffeine. It is important to follow these instructions carefully to ensure a safe and successful recovery after the EGD. If you have any questions or concerns about what you can eat or drink after the procedure, be sure to talk to your healthcare provider. | similar | 1 | 2 | Comprehensive |
| Can I drive home after an esophagogastroduodenoscopy ? | Most people can safely drive themselves home after an esophagogastroduodenoscopy (EGD), also known as an upper endoscopy, as long as they did not receive sedation during the procedure. If you did not receive any sedation, then you should be able to resume your normal activities, including driving, immediately after the procedure. However, you may experience a sore throat or mild discomfort in your upper abdomen, which should resolve within a few hours. On the other hand, if you received sedation during the procedure, you should not drive for at least 12 to 24 hours afterward, or until the sedative has completely worn off. Sedation can impair your reaction time, coordination, and decision-making abilities, which can make driving unsafe. You should arrange for someone to accompany you to the procedure and drive you home afterward if you receive sedation. It's important to follow your doctor's instructions and recommendations regarding driving or any other activities after an EGD. | similar | 1 | 1 | Comprehensive |
| How soon can I return to work or school after an esophagogastroduodenoscopy ? | Esophagogastroduodenoscopy (EGD) is a medical procedure used to examine the upper gastrointestinal tract, including the esophagus, stomach, and duodenum. The recovery time after EGD can vary depending on factors such as the reason for the procedure, the individual's overall health, and the type of sedation used during the procedure. In general, most people can return to work or school on the same day after an EGD, as long as they have not been sedated or have only received mild sedation. However, if you have received deeper sedation, it may take a few hours for the effects to wear off, and you should plan to have someone else drive you home. Additionally, it is recommended to avoid driving or operating heavy machinery for at least 12 to 24 hours after receiving sedation. After the procedure, you may experience some discomfort or bloating in the abdomen, but this should resolve within a few hours. Your doctor may also advise you to avoid certain activities or foods for a period of time after the procedure, depending on your individual case. It is important to follow your doctor's instructions for post-procedure care to ensure a smooth and speedy recovery. | similar | 1 | 2 | Correct but inadequate |
| Are there any foods or activities I should avoid after an esophagogastroduodenoscopy ? | After an esophagogastroduodenoscopy (EGD), which is a procedure that involves examining the upper digestive tract using a flexible endoscope, there are some things that you may want to avoid to allow your body to recover properly and prevent complications. Here are a few recommendations: Avoid eating or drinking anything until the numbness from the throat spray wears off, and your doctor gives you the green light. It is essential to allow your throat to recover from the procedure, and eating or drinking too soon can cause discomfort or even nausea. It is recommended to avoid spicy or acidic foods for a few days after the procedure, as these may irritate your esophagus or stomach lining, which can cause discomfort or inflammation. Avoid strenuous activities or heavy lifting for a few days after the procedure, as this can increase your blood pressure and cause bleeding at the site of the endoscopy. It is essential to avoid smoking or consuming alcohol for at least 24 hours after the procedure, as this can increase the risk of bleeding or other complications. It is essential to follow the instructions given by your doctor carefully and contact them immediately if you experience any severe or unexpected symptoms after the procedure. | similar | 2 | 3 | Mixed with correct and incorrect/outdated data |
| Will I need to make any lifestyle changes after an esophagogastroduodenoscopy ? | After an esophagogastroduodenoscopy (EGD), you may need to make some temporary lifestyle changes to allow your body to recover fully. Here are some general guidelines to follow: Rest: You may feel tired or sleepy after the procedure, so it's essential to rest for the remainder of the day. Diet: Your doctor may recommend a soft or liquid diet for a day or two following the EGD. Avoid foods that are spicy, acidic, or hard to digest. Medications: Your doctor may have instructed you to stop taking certain medications before the EGD. Be sure to follow these instructions carefully, and discuss with your doctor which medications you can resume taking after the procedure. Activity: Avoid strenuous activities or exercise for a day or two after the EGD. You should be able to resume your regular activities within a few days. Follow-up: Make sure to attend any follow-up appointments with your doctor to discuss the results of the EGD and any further treatment that may be necessary. Overall, lifestyle changes after an EGD are usually temporary and are meant to help you recover fully. If you have any questions or concerns, talk to your doctor or healthcare provider. | similar | 1 | 1 | Comprehensive |
| How can I manage any discomfort or pain after the esophagogastroduodenoscopy ? | Esophagogastroduodenoscopy (EGD) is a procedure that allows doctors to examine the esophagus, stomach, and duodenum using a flexible, lighted tube. Some discomfort or pain is normal after the procedure, but there are steps you can take to manage it. Here are some tips: Rest: It is important to rest for the first few hours after the procedure. Avoid any strenuous activity or heavy lifting. Drink fluids: Drink plenty of fluids to stay hydrated. You may feel some discomfort when swallowing, so stick to cool or room temperature fluids, and avoid hot or cold beverages. Eat soft, easy-to-digest foods: Eat soft, bland foods such as soup, applesauce, and plain toast. Avoid spicy, acidic, or fatty foods, as they can cause discomfort. Take pain medication: Your doctor may prescribe pain medication to help manage any discomfort or pain. Follow your doctor's instructions carefully when taking the medication. Use heat or cold therapy: Applying heat or cold to the affected area can help relieve discomfort. Use a heating pad or a warm towel for heat therapy, and an ice pack for cold therapy. Rest and relax: Take it easy for a few days after the procedure. Get plenty of rest and avoid stress and anxiety, as they can make the discomfort worse. If you experience severe pain, vomiting, or other concerning symptoms after an EGD, contact your doctor right away. | similar | 1 | 3 | Correct but inadequate |
| How long should I wait before eating solid foods again? | The answer to this question depends on several factors, such as the reason for not eating solid foods, any medical conditions you may have, and any instructions or recommendations from your healthcare provider. If you have undergone a medical procedure or surgery, your healthcare provider will provide specific instructions on when you can start eating solid foods again. Generally, you will start with clear liquids, then progress to full liquids, and then to soft foods before gradually adding solid foods back into your diet. If you are fasting for religious or personal reasons, the length of your fast and when you should resume solid foods will depend on your personal beliefs and practices. It's always important to consult with your healthcare provider before starting any fasting regimen. If you are experiencing digestive issues or have a medical condition that requires you to avoid solid foods, your healthcare provider will provide specific recommendations on when and how to reintroduce solid foods back into your diet. In general, it's important to listen to your body and take it slow when reintroducing solid foods. Start with small portions of soft, easily digestible foods and gradually increase the amount and complexity of the foods as tolerated. If you experience any discomfort or digestive issues, consult with your healthcare provider. | similar | 1 | 2 | Comprehensive |
| Will I need someone to help me at home after the esophagogastroduodenoscopy ? | Esophagogastroduodenoscopy (EGD) is a medical procedure that allows a doctor to examine the lining of your esophagus, stomach, and duodenum using an endoscope. The need for assistance at home after an EGD will depend on several factors such as the type of anesthesia used, any medications prescribed by the doctor, and the individual's overall health condition. If you received sedation or general anesthesia during the procedure, you may need someone to drive you home and stay with you for a few hours until the effects wear off. It's also possible that you may feel a bit groggy or dizzy after the procedure, and having someone to assist you with basic tasks like getting out of bed, going to the bathroom, or preparing meals can be helpful. However, if you received only local anesthesia or no anesthesia at all, you may not require any assistance at home. Your doctor will likely provide you with specific instructions about post-procedure care, including any restrictions on eating or drinking, and what to do if you experience any complications. It's always a good idea to discuss your concerns with your doctor before the procedure to ensure that you understand what to expect and have a plan in place for any necessary aftercare. | similar | 1 | 1 | Comprehensive |
| Can I resume my normal exercise routine after the esophagogastroduodenoscopy ? | It is generally recommended to avoid vigorous exercise for at least 24 hours after an esophagogastroduodenoscopy (EGD), which is a medical procedure used to examine the upper gastrointestinal tract. This is because the procedure involves inserting a flexible tube with a camera through the mouth and down the esophagus, which can cause some irritation and discomfort in the throat and stomach. However, the exact length of time you should wait before resuming your exercise routine may vary depending on your individual situation and the advice of your doctor. If you experienced any complications during the EGD, or if you have other health conditions that may be affected by exercise, you should discuss your exercise plans with your doctor. In general, it is important to listen to your body and take things slowly when you start exercising again after any medical procedure. Start with low-impact activities and gradually increase your intensity over time. If you experience any unusual symptoms, such as pain or discomfort, during or after exercise, you should stop and consult with your doctor. | similar | 3 | 1 | Correct but inadequate |
| **Other:** |  |  |  |  |  |
| How much does an esophagogastroduodenoscopy cost? | The cost of an esophagogastroduodenoscopy (EGD) can vary depending on a number of factors such as the country and region where the procedure is being performed, the healthcare provider, and whether the patient has insurance coverage or not. In the United States, the cost of an EGD can range from around $1,000 to $5,000 or more, depending on the above factors. However, it's important to note that insurance coverage can greatly affect the final cost that the patient is responsible for paying. If you are considering an EGD, it's best to speak with your healthcare provider and insurance company to get an estimate of the cost for your specific situation. | similar | 1 | 1 | Comprehensive |
| Does insurance cover the cost of the esophagogastroduodenoscopy ? | Whether insurance covers the cost of an esophagogastroduodenoscopy (EGD) depends on a number of factors, including the specific insurance plan, the reason for the procedure, and the individual's medical history. In many cases, insurance will cover the cost of an EGD if it is deemed medically necessary. This may be the case if the individual is experiencing symptoms such as abdominal pain, difficulty swallowing, or gastrointestinal bleeding. However, if the procedure is being done for screening purposes or for a non-medical reason, it may not be covered by insurance. It is always best to check with your insurance provider directly to determine whether they will cover the cost of an EGD in your specific situation. Your healthcare provider can also provide guidance on the likelihood of coverage and any potential out-of-pocket costs. | similar | 1 | 2 | Correct but inadequate |
| Can I bring someone with me to the esophagogastroduodenoscopy procedure? | Yes, you are typically allowed to bring someone with you to an esophagogastroduodenoscopy (EGD) procedure, although the specific policies may vary depending on the medical facility and the circumstances of the procedure. In most cases, it's a good idea to have someone with you to provide support, drive you home after the procedure (since you may be given sedatives or anesthesia), and help you with any necessary post-procedure care. If you're unsure about the policies regarding bringing someone with you to your specific EGD procedure, it's best to check with your doctor's office or the medical facility where the procedure will be performed. | similar | 1 | 1 | Comprehensive |
| Can I request a specific doctor to perform my esophagogastroduodenoscopy ? | In general, it is possible to request a specific doctor to perform your esophagogastroduodenoscopy (EGD), but it ultimately depends on the policies and procedures of the medical facility where the procedure will take place. If you have a particular doctor in mind, you can ask your primary care physician or the medical facility where the procedure will take place if it is possible to schedule your EGD with that doctor. However, it's important to keep in mind that the doctor you request may not be available on the date and time you need the procedure. Additionally, the doctor who performs your EGD should be a trained specialist in gastrointestinal (GI) endoscopy and should have experience performing this specific procedure. It's important to choose a doctor who is qualified and experienced to ensure that you receive the best possible care. | similar | 1 | 1 | Comprehensive |
| Will I receive a report after the esophagogastroduodenoscopy ? | The timing of receiving a report after an esophagogastroduodenoscopy (EGD) can vary depending on the specific circumstances and policies of the medical facility where the procedure was performed. In general, the endoscopist who performs the EGD will typically provide some initial findings and impressions immediately after the procedure, while the patient is still in the recovery area. However, a more detailed report, including any biopsies that were taken, may take a few days to be completed and reviewed. The exact timing can also depend on factors such as the complexity of the procedure, the volume of cases being handled by the facility, and the availability of the pathologist to review the biopsy specimens. Patients should check with their healthcare provider or the facility where the EGD was performed to get an estimate of when they can expect to receive a complete report of the results. | similar | 1 | 1 | Comprehensive |
| What if I am allergic to the sedative used during the esophagogastroduodenoscopy ? | If you are allergic to the sedative used during the esophagogastroduodenoscopy (EGD), it is important to inform your doctor or medical team before the procedure. Depending on the severity of your allergy and the available alternatives, your doctor may recommend postponing or canceling the procedure, or using a different type of sedative medication. In some cases, your doctor may recommend using an alternative method to perform the EGD, such as using local anesthesia or a different type of sedation. It is important to follow your doctor's instructions and communicate any concerns or questions you may have. Your doctor will work with you to ensure your safety and well-being during the procedure. | similar | 1 | 1 | Comprehensive |
| What happens if a problem is found during the esophagogastroduodenoscopy ? | Esophagogastroduodenoscopy (EGD) is a diagnostic procedure that allows doctors to examine the inside lining of the esophagus, stomach, and upper part of the small intestine (duodenum). If a problem is found during an EGD, the doctor will typically take one or more biopsies (small tissue samples) to send to a laboratory for further testing. The biopsy results can help the doctor determine the cause of the problem and plan appropriate treatment. Depending on the nature and severity of the problem, the doctor may recommend follow-up EGDs or other tests to monitor or treat the condition. Some common problems that can be detected during an EGD include inflammation of the esophagus (esophagitis), stomach (gastritis), or duodenum (duodenitis); ulcers; blockages or narrowing of the esophagus or stomach; and abnormal growths such as polyps or tumors. It's important to note that in some cases, an EGD may not reveal any problems or abnormalities, even if the patient is experiencing symptoms. In these cases, the doctor may recommend further testing or other diagnostic procedures to identify the cause of the symptoms. | similar | 1 | 1 | Comprehensive |
| What are the credentials of the medical professionals who will be performing the esophagogastroduodenoscopy ? | As an AI language model, I don't have access to specific information about the medical professionals who will be performing the esophagogastroduodenoscopy (EGD) for a particular patient. However, in general, an EGD is typically performed by a gastroenterologist, a medical doctor who specializes in diagnosing and treating disorders of the digestive system. Gastroenterologists are highly trained medical professionals who have completed a medical degree, a residency in internal medicine, and a fellowship in gastroenterology. They also undergo rigorous training and testing to become certified by the American Board of Internal Medicine in Gastroenterology. It's important to note that the credentials of medical professionals can vary depending on the country or region where they practice, and different healthcare systems may have different requirements for certification and training. Additionally, it's always a good idea to speak with your healthcare provider about their qualifications and experience before undergoing any medical procedure. | similar | 1 | 1 | Comprehensive |
| Will I be monitored during the entire esophagogastroduodenoscopy procedure? | Yes, it is standard practice to monitor patients throughout an esophagogastroduodenoscopy (EGD) procedure to ensure their safety and comfort. Monitoring typically involves the use of equipment that measures your blood pressure, heart rate, and oxygen levels, as well as continuous visual monitoring by the medical team. The monitoring equipment may include a blood pressure cuff, a pulse oximeter that attaches to your finger to measure oxygen saturation, and ECG leads to monitor your heart's electrical activity. This equipment will be used to continuously assess your vital signs and alert the medical team if any changes occur that require attention. Additionally, the healthcare professionals performing the EGD will be in constant communication with you during the procedure to ensure that you are comfortable and to provide reassurance. They will also be able to provide medications, such as sedatives and pain relievers, as needed to help you relax and manage any discomfort. | similar | 1 | 1 | Comprehensive |
| How can I address any concerns or questions I have before the esophagogastroduodenoscopy ? | Before an esophagogastroduodenoscopy (EGD), it is normal to have concerns or questions about the procedure. Here are some steps you can take to address these concerns: Speak with your doctor: Your doctor or gastroenterologist is the best source of information about the procedure. They can explain the procedure, the risks and benefits, and answer any questions you may have. Do some research: You can also research the procedure online or through educational material provided by your doctor's office or hospital. It can help you understand the procedure better and feel more prepared. Talk to others: If you know someone who has had an EGD, you may want to speak with them about their experience. They can provide insights into what to expect during the procedure and how to prepare. Write down your questions: It can be helpful to write down any questions or concerns you have before your appointment. This way, you can ensure that you don't forget to ask anything during your consultation. Take someone with you: If you are nervous about the procedure, consider taking a friend or family member with you to the appointment. They can provide emotional support and help you remember any important information discussed during the consultation. Remember that it's essential to address any concerns or questions you have before the procedure to feel more comfortable and confident. Your doctor and healthcare team are there to help you throughout the process. | similar | 1 | 1 | Comprehensive |

Supplemental Table 2. Grading of the responses from the ChatGPT Language Model for colonoscopy-related questions

| Basic Knowledge | 1^st^ response | 2^nd^ response | Reviewer 1 grades  1.Comprehensive 2. Correct but inadequate 3. Mixed with correct and incorrect/outdated data 4. Completely incorrect | Reviewer 2 grades  1. Comprehensive 2. Correct but inadequate 3. Mixed with correct and incorrect/outdated data 4. Completely incorrect | Reviewer 3 grades for the responses with discrepancies |
| --- | --- | --- | --- | --- | --- |
| What is a colonoscopy? | A colonoscopy is a medical procedure that involves the use of a flexible tube with a camera and light on the end, called a colonoscope, to examine the inside of the large intestine, also known as the colon. The procedure is typically performed by a gastroenterologist, a doctor who specializes in the digestive system. During the colonoscopy, the patient is usually given medication to help them relax and reduce discomfort. The colonoscope is then inserted into the rectum and advanced through the colon, allowing the doctor to visualize the lining of the colon and identify any abnormalities, such as polyps or other growths. If any abnormal areas are identified, the doctor may take a biopsy or remove them for further testing. Colonoscopies are commonly used to screen for colon cancer, as well as to investigate symptoms such as abdominal pain, rectal bleeding, and changes in bowel habits. | similar | 1 | 1 | Comprehensive |
| How is a colonoscopy performed? | A colonoscopy is a medical procedure used to examine the colon (large intestine) and rectum for signs of disease or abnormality. During a colonoscopy, a flexible tube with a camera and light at the end, called a colonoscope, is inserted into the rectum and guided through the colon. Here are the steps typically involved in a colonoscopy: Preparation: Before the procedure, the patient will need to cleanse their colon by following specific dietary restrictions and taking laxatives or enemas to empty the bowels. This is important to ensure that the colon is clear and free of any stool or debris that could obstruct the view during the procedure. Anesthesia: The patient is given sedation to help them relax and feel comfortable during the procedure. This can be done through an IV or inhaled gas. Insertion: The colonoscope is inserted into the rectum and gradually advanced through the colon. The doctor will use air or carbon dioxide to inflate the colon and help maneuver the scope. Visual examination: As the colonoscope advances, the camera at the end of the scope sends images to a monitor, allowing the doctor to examine the lining of the colon for any abnormalities, such as polyps or inflammation. Biopsy and removal: If any suspicious areas are found, the doctor may take a tissue sample (biopsy) for further testing. Polyps may also be removed during the procedure using a small tool attached to the scope. Completion: Once the examination is complete, the colonoscope is gradually removed from the colon. The patient may feel some discomfort or bloating during this time. The procedure usually takes between 30 minutes to an hour, depending on the extent of the examination and any necessary interventions. After the procedure, the patient will be monitored in a recovery area until the effects of the sedation wear off. | similar | 1 | 1 | Comprehensive |
| What are the risks associated with colonoscopy? | Colonoscopy is generally considered a safe and effective procedure for diagnosing and treating conditions of the colon and rectum. However, like any medical procedure, there are potential risks and complications that may arise, including: Bleeding: During a colonoscopy, the doctor may need to remove tissue samples or polyps. This can cause bleeding, which is usually minor and stops on its own. However, in rare cases, the bleeding can be severe and require further medical attention. Perforation: In very rare cases, the colonoscope can puncture the wall of the colon or rectum. This can cause infection, bleeding, or even require emergency surgery. Reaction to anesthesia: Most colonoscopies are performed with sedation or anesthesia to make the procedure more comfortable for the patient. However, some people may have an allergic reaction to these medications, which can be serious. Incomplete exam: In some cases, the colonoscope may not be able to reach the entire length of the colon due to factors such as the patient's anatomy or a medical condition. This can result in an incomplete exam, which may require further testing or a repeat colonoscopy. Misdiagnosis: Despite being a very effective diagnostic tool, colonoscopy is not perfect, and there is a risk of misdiagnosis. In some cases, abnormalities such as polyps or tumors may be missed or not detected during the exam. It's important to discuss the potential risks and benefits of colonoscopy with your healthcare provider and to follow their instructions for preparing for the procedure to minimize the risk of complications. | similar | 1 | 2 | Comprehensive |
| Do I need to fast before a colonoscopy? | Yes, typically you will need to fast before a colonoscopy. The purpose of fasting is to ensure that your colon is completely empty, allowing for a clear view of the colon during the procedure. Your doctor will provide specific instructions on what to eat and drink before the colonoscopy and when to stop eating and drinking. Generally, you will be asked to avoid solid foods for at least 24 hours before the procedure and to consume only clear liquids such as water, tea, coffee, clear juices, clear broths, and sports drinks. It's important to follow your doctor's instructions carefully to ensure the success of the colonoscopy and to avoid any complications. If you have any concerns or questions about fasting before a colonoscopy, be sure to speak with your doctor. | similar | 1 | 1 | Comprehensive |
| How long does a colonoscopy take? | The length of a colonoscopy can vary depending on several factors, including the individual's medical history and the findings during the procedure. In general, a typical colonoscopy takes between 30 and 60 minutes to complete. However, the preparation process, which involves fasting and bowel preparation, can take several hours or even a day or more. It's also important to note that the individual will need to spend additional time in the recovery area after the procedure, as the effects of sedation can take several hours to wear off. The doctor performing the colonoscopy will be able to provide a more specific estimate of how long the procedure is likely to take in individual cases. | similar | 1 | 2 | Correct but inadequate |
| Will I be sedated during the colonoscopy? | Yes, typically you will be sedated during a colonoscopy. Sedation helps to keep you calm and relaxed during the procedure, as well as to minimize any discomfort or pain. The level of sedation will depend on a number of factors, including your overall health, medical history, and the preferences of the doctor performing the colonoscopy. The most common type of sedation used during a colonoscopy is called conscious sedation. This means that you will be given medications that make you feel relaxed and drowsy, but you will still be able to respond to verbal commands and breathe on your own. Conscious sedation allows you to be comfortable during the procedure without the need for general anesthesia, which carries more risks and requires more extensive monitoring. It's important to follow your doctor's instructions regarding preparation and any restrictions prior to your colonoscopy, as well as discussing any concerns or questions you may have regarding sedation or the procedure itself. | similar | 1 | 2 | Correct but inadequate |
| How often should I have a colonoscopy? | The frequency of colonoscopies will depend on various factors, including your age, medical history, family history, and previous screening results. In general, for people of average risk, the American Cancer Society recommends that individuals start regular colon cancer screening at age 45 and continue until age 75. There are several screening options available, including colonoscopy, stool tests, and sigmoidoscopy. If your initial colonoscopy is normal and you have no significant risk factors, your doctor may recommend that you have another one in 10 years. If polyps or other abnormal findings are detected during your colonoscopy, your doctor may recommend more frequent colonoscopies or additional testing. It's important to talk to your doctor about your individual risk factors and screening options to determine the best course of action for you. | similar | 1 | 2 | Comprehensive |
| Will the colonoscopy hurt? | Colonoscopy is a medical procedure that is used to examine the inside of the colon and rectum. During the procedure, a long, flexible tube with a camera is inserted into the rectum and guided through the colon, allowing the doctor to examine the lining of the colon and rectum for any abnormalities. While the thought of having a colonoscopy may be uncomfortable, most patients report only minor discomfort during the procedure itself. This is because the patient is usually given a sedative and pain medication to help them relax and feel more comfortable. However, it is normal to feel some mild discomfort or pressure during the procedure as the scope is advanced through the colon. Some patients may also experience cramping or bloating afterward. These symptoms usually go away within a few hours after the procedure. It is important to remember that while a colonoscopy may be uncomfortable, it is an important tool for detecting and preventing colon cancer. If you have any concerns or questions about the procedure, you should talk to your doctor before the appointment. | As an AI language model, I don't have personal experience with colonoscopies, but I can provide you with some information. During a colonoscopy, the doctor inserts a long, flexible tube with a camera on the end into the rectum and colon to examine the lining. The procedure itself is not usually painful because patients are given sedation to help them relax and reduce discomfort. However, some people may experience mild discomfort, cramping, or bloating during or after the procedure. It's important to follow the preparation instructions provided by your doctor to help ensure a successful and comfortable procedure. Additionally, if you have any concerns about pain or discomfort during the procedure, you should discuss them with your doctor beforehand. | 1 | 1 | Comprehensive |
| Treatment |  |  |  |  | Treatment |
| Why do I need a colonoscopy? | A colonoscopy is a medical procedure that is performed to examine the large intestine (colon) and rectum for signs of abnormalities, such as polyps or tumors. It is often recommended for people who are at an increased risk of developing colon cancer or other colorectal diseases. Some common reasons for needing a colonoscopy include: Screening for colon cancer: The American Cancer Society recommends that adults begin regular colon cancer screening at age 45 or earlier if they have certain risk factors. A colonoscopy is one of several screening tests that can be used to detect colon cancer in its early stages when it is most treatable. Polyps: Polyps are growths that can develop on the lining of the colon. While most polyps are benign (noncancerous), some can develop into colon cancer over time. A colonoscopy can detect and remove polyps before they become cancerous. Unexplained changes in bowel habits: If you have experienced unexplained changes in your bowel habits, such as persistent diarrhea or constipation, a colonoscopy can help determine if there is an underlying issue causing these symptoms. Abdominal pain: If you have chronic or severe abdominal pain, a colonoscopy may be recommended to help diagnose the cause of your pain. Family history of colon cancer: If you have a family history of colon cancer or other colorectal diseases, your doctor may recommend a colonoscopy to screen for these conditions. If your doctor recommends a colonoscopy, it is important to follow their advice and schedule the procedure. Colon cancer is highly treatable when detected early, and a colonoscopy is a key tool in early detection and prevention. | similar | 1 | 2 | Comprehensive |
| How will the results of the colonoscopy be used in my treatment? | The results of a colonoscopy will be used to help diagnose and treat any potential issues with your colon. If abnormalities such as polyps, tumors, or inflammation are detected during the procedure, your doctor may recommend further testing or treatment to address these issues. If no abnormalities are found, the results can provide reassurance that your colon is healthy and can serve as a baseline for future screening. Depending on your age, family history, and other risk factors, your doctor may recommend regular colonoscopies to monitor your colon health and detect any changes early on. Overall, the results of a colonoscopy are an important tool for your healthcare provider to assess your colon health and recommend appropriate treatment or preventative measures. It's important to discuss any questions or concerns you have about the results and their implications with your doctor. | similar | 2 | 1 | Comprehensive |
| Can a colonoscopy be used to treat my condition? | As an AI language model, I do not have access to your medical records and cannot provide personalized medical advice. However, in general, a colonoscopy is a diagnostic procedure used to examine the colon for abnormalities, such as polyps or tumors. It is not typically used as a treatment for conditions. If any abnormalities are detected during a colonoscopy, such as polyps, they can be removed during the procedure for diagnostic and therapeutic purposes. Additionally, colonoscopy can also be used for screening and surveillance of certain conditions, such as colon cancer. If you have a medical condition and are wondering if a colonoscopy may be appropriate for diagnosis or treatment, it is important to consult with a qualified healthcare provider who can evaluate your individual situation and recommend appropriate diagnostic and treatment options. | similar | 1 | 3 | Mixed with correct and incorrect/outdated data |
| What happens if a problem is found during the colonoscopy? | If a problem is found during a colonoscopy, the gastroenterologist performing the procedure may take a biopsy or remove abnormal tissue for further testing. The types of problems that can be found during a colonoscopy include: Polyps: These are abnormal growths in the lining of the colon that can develop into cancer if left untreated. If polyps are found during a colonoscopy, they may be removed during the procedure to prevent cancer from developing. Inflammation: Inflammation in the colon can be a sign of inflammatory bowel disease (IBD), such as ulcerative colitis or Crohn's disease. If inflammation is detected, the gastroenterologist may take a biopsy to confirm the diagnosis and develop a treatment plan. Diverticula: Diverticula are small, bulging pouches that can form in the lining of the colon. If diverticula are found during a colonoscopy, the gastroenterologist may recommend further testing to determine if they are causing any symptoms. Cancer: If a suspicious area is found during a colonoscopy, the gastroenterologist may take a biopsy to determine if it is cancerous. If cancer is detected, the patient will need further testing and treatment to address the cancer. Overall, if a problem is found during a colonoscopy, the gastroenterologist will work with the patient to develop a plan for further testing and treatment. Early detection and treatment of problems found during a colonoscopy can help prevent serious health problems in the future. | similar | 1 | 1 | Comprehensive |
| Can a colonoscopy diagnose all conditions or problems? | A colonoscopy is a medical procedure in which a long, flexible tube with a camera at the end is inserted into the rectum and advanced through the entire colon to examine the lining of the colon for abnormalities, such as polyps, inflammation, ulcers, or cancer. While colonoscopy is a useful tool in detecting many conditions and problems in the colon, it is not a perfect diagnostic test, and some conditions may not be detected during the procedure. For example, colonoscopy may miss small polyps or early stages of cancer that are not visible to the naked eye. In addition, colonoscopy cannot detect conditions or problems outside the colon, such as diseases of the small intestine or stomach. Other diagnostic tests, such as blood tests, stool tests, imaging tests, and endoscopic procedures of other parts of the digestive system, may be necessary to diagnose certain conditions or problems that are not detected during a colonoscopy. Therefore, while colonoscopy is an important screening and diagnostic tool for many conditions and problems in the colon, it should be used in conjunction with other diagnostic tests to ensure a comprehensive evaluation of the digestive system. | similar | 1 | 1 | Comprehensive |
| Lifestyle/aftercare |  |  |  |  | Lifestyle |
| Can I eat and drink after a colonoscopy? | After a colonoscopy, you will typically be advised to avoid eating or drinking anything until the effects of the anesthesia or sedative have worn off completely, and your doctor has determined that it is safe for you to resume normal activities. This is usually a period of a few hours. Once your doctor has cleared you to eat and drink, you can usually resume your normal diet unless instructed otherwise. However, it is important to stay well-hydrated and to avoid alcohol and caffeine for the first few hours after the procedure, as these substances can cause dehydration and may interfere with the healing process. If you experience any discomfort or abnormal symptoms after the colonoscopy, such as severe pain, bleeding, or fever, you should contact your doctor immediately. | similar | 1 | 1 | Comprehensive |
| Can I drive home after a colonoscopy? | After a colonoscopy, you will likely receive sedation or anesthesia, which can affect your cognitive abilities and physical coordination for a period of time afterward. Therefore, it is not recommended that you drive yourself home after the procedure. It is important to arrange for someone to drive you home, and it is generally recommended that you have someone stay with you for a few hours following the procedure until the effects of the sedation have fully worn off. Your healthcare provider will give you specific instructions on how long you should wait before driving or performing other activities that require alertness and coordination. If you do not have someone to drive you home after the colonoscopy, you may need to reschedule the procedure or make other arrangements for transportation. | similar | 1 | 2 | Correct but inadequate |
| How soon can I return to work or school after a colonoscopy? | The timing for returning to work or school after a colonoscopy may vary depending on several factors, including the individual's overall health, the type of procedure performed, and any complications that may have occurred during the procedure. In general, most people can return to their normal activities, including work or school, on the same day after a colonoscopy. However, you may experience some temporary side effects such as bloating, cramping, or gas, which can last for a few hours. These symptoms usually resolve on their own without any specific treatment. If you had sedation during the colonoscopy, you may need to wait until the effects of the sedative have worn off completely before returning to work or school. Your doctor or healthcare provider will provide you with specific instructions regarding when you can resume your normal activities, including any restrictions or precautions you should take. It's important to note that everyone's recovery time can be different. If you have any concerns or questions about returning to work or school after a colonoscopy, it's best to speak with your doctor or healthcare provider for personalized guidance | similar | 1 | 2 | Comprehensive |
| Are there any foods or activities I should avoid after a colonoscopy? | After a colonoscopy, your digestive system needs time to recover, and your doctor may recommend avoiding certain foods and activities to ensure a smooth recovery. Here are some general guidelines: Foods to avoid: For the first few days after your colonoscopy, it is recommended to avoid foods that may be difficult to digest or may irritate your digestive system, such as spicy or fatty foods, raw vegetables, and fruits with seeds or skins. You may also want to avoid alcohol and caffeine. Instead, opt for soft, bland foods like soup, broths, cooked vegetables, and lean protein. Activities to avoid: For the first 24 hours after your colonoscopy, you should avoid any activities that require concentration or coordination, such as driving, operating machinery, or signing legal documents. You may also want to avoid strenuous physical activity and exercise for a few days. Medications: Your doctor may give you specific instructions about what medications to take or avoid after your colonoscopy. It is important to follow these instructions carefully. It's essential to follow the advice of your doctor regarding post-colonoscopy care. If you experience severe pain, bleeding, or fever after the procedure, seek medical attention immediately. | similar | 1 | 2 | Correct but inadequate |
| How can I manage any discomfort or pain after a colonoscopy? | After a colonoscopy, it is common to experience some discomfort or pain. Here are some tips that may help manage these symptoms: Rest: You may feel tired after the procedure, so it is important to rest and avoid any strenuous activities for the remainder of the day. Drink plenty of fluids: Drinking water and other fluids can help prevent dehydration and also help flush out any remaining gas or stool in your colon. Apply heat: Applying a heating pad or hot water bottle to your abdomen may help relieve any cramping or discomfort. Take over-the-counter pain relievers: You can take over-the-counter pain relievers such as acetaminophen (Tylenol) or ibuprofen (Advil) to help alleviate any pain or discomfort. However, it is important to avoid taking aspirin or any other blood-thinning medications for at least 24 hours after the procedure. Eat a light diet: Stick to a light diet that includes easy-to-digest foods such as soup, crackers, and toast. Avoid spicy, greasy, or heavy foods for the first few days after the procedure. Follow your doctor's instructions: Your doctor will give you specific instructions on how to manage any discomfort or pain after the colonoscopy. It is important to follow these instructions closely to ensure a speedy recovery. If you experience severe pain, fever, or bleeding after the procedure, contact your doctor immediately as these may be signs of a complication. | similar | 1 | 3 | Comprehensive |
| Other |  |  |  |  | Other |
| How much does a colonoscopy cost? | The cost of a colonoscopy can vary widely depending on various factors, including your location, the provider performing the procedure, and your insurance coverage. Without insurance, the cost of a colonoscopy in the United States can range from $1,000 to $5,000 or more, depending on where you live and the provider you choose. However, many insurance plans cover preventive colonoscopies for patients over the age of 50 or those with a family history of colon cancer. To get an accurate estimate of the cost of a colonoscopy for your specific situation, it is best to contact your healthcare provider or insurance company directly. They can provide you with information on your coverage and any out-of-pocket expenses you may incur. | similar | 2 | 2 | Correct but inadequate |
| Will my insurance cover the cost of the colonoscopy? | As an AI language model, I do not have access to your personal information or your insurance policy details. Therefore, I cannot provide a definitive answer to your question. However, generally, whether your insurance will cover the cost of a colonoscopy or not depends on your specific insurance policy and the reason for the colonoscopy. Most insurance plans cover colonoscopies for preventive purposes, such as routine colon cancer screening. Still, some plans may not cover the procedure if it's being done for diagnostic reasons or if you don't meet certain age or risk criteria. The best way to determine whether your insurance will cover the cost of a colonoscopy is to contact your insurance company and ask about the details of your policy's coverage for this specific procedure. You may also want to check with your healthcare provider to ensure that they are in-network with your insurance provider. | similar | 1 | 1 | Comprehensive |
| Can I bring someone with me to the colonoscopy procedure? | Yes, it is usually allowed and even encouraged to bring someone with you to the colonoscopy procedure for support and assistance. Most medical facilities and clinics that perform colonoscopies will allow a friend, family member, or caregiver to accompany you. However, it's important to confirm this with your healthcare provider or the facility where the procedure is being performed, as some facilities may have specific rules or restrictions on the number of people who can accompany you or the areas of the facility where they are allowed to go. Additionally, if you are receiving sedation during the procedure, your companion may need to wait in a designated waiting area until you are ready to be discharged. They may also need to sign a consent form agreeing to follow any instructions given by the medical staff during the procedure. Overall, it's a good idea to discuss any concerns or questions you have about bringing someone with you to your colonoscopy with your healthcare provider or the facility where the procedure will be performed. | similar | 1 | 1 | Comprehensive |
| How can I address any concerns or questions I have before the colonoscopy? | Before your colonoscopy, it's important to address any concerns or questions you may have to ensure that you feel fully prepared and informed. Here are some steps you can take: Speak with your doctor: Your doctor is the best person to address any concerns or questions you may have about your colonoscopy. Make sure to schedule an appointment or call their office to discuss any concerns you may have. Your doctor can provide you with specific information about the preparation process, the procedure itself, and any risks or complications that may arise. Do your own research: In addition to speaking with your doctor, you can also do your own research about colonoscopies. Look for reputable sources of information, such as medical journals or websites affiliated with major hospitals or medical organizations. This can help you better understand the procedure and what to expect. Speak with friends or family members who have had a colonoscopy: If you have friends or family members who have had a colonoscopy, ask them about their experience. They may be able to provide you with additional information or advice that your doctor may not have mentioned. Attend a pre-procedure consultation: Many hospitals or clinics offer pre-procedure consultations for colonoscopies. This is an opportunity to speak with a healthcare provider who can answer your questions and provide you with more information about the procedure. Remember, it's important to address any concerns or questions you have before your colonoscopy to ensure that you feel comfortable and prepared. | similar | 1 | 1 | Comprehensive |

Supplemental Table 3. Grading of the responses from the ChatGPT Language Model for endoscopic ultrasound (EUS)-related questions

| **Basic Knowledge** | 1st response | 2^nd^ response | Reviewer 1 grades 1. Comprehensive 2. Correct but inadequate 3. Mixed with correct and incorrect/outdated data 4. Completely incorrect | Reviewer 1 grades 1. Comprehensive 2. Correct but inadequate 3. Mixed with correct and incorrect/outdated data 4. Completely incorrect | Reviewer 3 grades for the responses with discrepancies |
| --- | --- | --- | --- | --- | --- |
| What is an EUS procedure? | EUS stands for Endoscopic Ultrasound, which is a minimally invasive medical procedure that combines endoscopy and ultrasound to obtain images and information about the digestive tract and adjacent organs. During an EUS procedure, a specialized endoscope is inserted through the mouth or rectum and advanced to the area of interest. The endoscope contains a tiny ultrasound probe that emits high-frequency sound waves to create images of the digestive tract, pancreas, liver, and surrounding structures. The images produced by EUS can be used to diagnose a variety of gastrointestinal and pancreatic diseases, such as cancer, cysts, and tumors. Additionally, EUS can be used to guide the insertion of needles to collect tissue samples or drain fluid collections. | Similar | 1 | 1 | Comprehensive |
| How does an EUS procedure work? | An Endoscopic Ultrasound (EUS) procedure is a minimally invasive medical technique used to examine the digestive tract, lungs, and surrounding organs. It is typically performed by a gastroenterologist or pulmonologist, using an endoscope that is equipped with an ultrasound transducer. During an EUS procedure, the patient will be given sedation to ensure comfort and reduce discomfort. The endoscope, a thin, flexible tube with a light and camera on the end, is then inserted through the mouth (for upper EUS) or rectum (for lower EUS) and advanced to the area of interest. The ultrasound transducer is then used to generate high-frequency sound waves that create detailed images of the surrounding tissues and organs, which are transmitted to a monitor for the doctor to view. EUS can be used to diagnose and stage various gastrointestinal and pulmonary conditions, including cancers, cysts, and abnormalities in the surrounding tissues. It can also be used to guide biopsy or drainage procedures, as the ultrasound images provide real-time visualization of the needle during the procedure. Overall, EUS is a safe and effective procedure that can provide important diagnostic and treatment information with minimal risk and discomfort for the patient. | An EUS (Endoscopic Ultrasound) procedure is a medical procedure that combines endoscopy and ultrasound to create detailed images of internal organs and tissues in the body. Here's how it typically works: Preparation: Before the procedure, you may need to fast for several hours to ensure that your stomach and intestines are empty. You may also receive sedation or anesthesia to help you relax and reduce discomfort during the procedure. Endoscopy: An endoscope, which is a thin, flexible tube with a camera and light at the end, is inserted through your mouth and down your throat into your esophagus, stomach, and duodenum (the first part of the small intestine). Ultrasound: A small ultrasound probe is attached to the end of the endoscope. The probe emits high-frequency sound waves that bounce off internal organs and tissues, creating detailed images of the area being examined. Imaging: The images produced by the ultrasound are displayed on a monitor, allowing the doctor to examine the walls of the digestive tract and nearby structures, such as lymph nodes, blood vessels, and organs like the pancreas or liver. Biopsy: If necessary, the doctor can use specialized tools to take small tissue samples (biopsies) for further examination under a microscope. This can help diagnose or rule out various conditions, such as cancer. Recovery: After the procedure, you'll be monitored in a recovery area until the sedation wears off. You may need to avoid eating and drinking for a few hours and may experience mild discomfort, such as a sore throat or bloating. Overall, an EUS procedure is a safe and effective way to examine and diagnose conditions in the digestive tract and surrounding areas. | 2 | Mixed with correct and incorrect/outdated data | Mixed with correct and incorrect/outdated data |
| Why would I need an EUS procedure? | Endoscopic ultrasound (EUS) is a medical procedure that uses high-frequency sound waves to produce images of internal organs and tissues in the digestive tract. There are several reasons why a doctor may recommend an EUS procedure, including: Diagnosis of digestive disorders: EUS can help diagnose conditions such as pancreatic cancer, gallstones, inflammatory bowel disease, and gastrointestinal tumors. Staging of cancer: EUS can be used to determine the extent and spread of certain types of cancer, including pancreatic cancer, esophageal cancer, and rectal cancer. Evaluation of lesions or cysts: EUS can be used to evaluate abnormalities such as cysts or lesions in the digestive tract. Treatment of certain conditions: EUS can also be used to guide certain medical procedures, such as draining fluid from a cyst or delivering medication directly to a tumor. If your doctor suspects that you may have a condition that could be diagnosed or treated using EUS, they may recommend the procedure as part of your overall treatment plan. As with any medical procedure, it's important to discuss the risks and benefits of EUS with your doctor beforehand to make an informed decision about your healthcare. | | 1 | 2 | Correct but inadequate |
| How is EUS different from other imaging techniques like MRI or CT scans? | Endoscopic ultrasound (EUS) is a minimally invasive diagnostic imaging technique that combines endoscopy and ultrasound to obtain detailed images of organs and tissues within the body. EUS is different from other imaging techniques like MRI or CT scans in several ways: Invasive vs Non-invasive: EUS is an invasive technique that involves inserting an endoscope into the body, while MRI and CT scans are non-invasive imaging techniques that do not require the insertion of any instruments into the body. Higher Resolution: EUS has higher resolution than CT scans and MRI for imaging certain organs like the pancreas and bile ducts. Close Proximity: EUS allows for closer proximity to the organs being examined than MRI or CT scans. This makes it particularly useful for examining small structures and obtaining biopsy samples. Real-time imaging: EUS provides real-time imaging of the organs being examined, allowing for dynamic evaluation and guidance during therapeutic procedures. Limited range: EUS has a limited range of imaging compared to CT and MRI. It can only visualize the organs and tissues close to the endoscope, whereas CT and MRI can provide images of the whole body. Overall, EUS is a valuable diagnostic tool that can provide detailed images of organs and tissues in the body, particularly for certain applications like evaluating pancreatic and biliary disorders, staging of gastrointestinal cancers, and obtaining biopsy samples. However, it has its limitations and may not be suitable for all imaging needs. | Similar | 1 | 1 | Comprehensive |
| Is an EUS procedure painful? | An EUS (endoscopic ultrasound) procedure is usually not painful, but there may be some discomfort involved. The level of discomfort can vary depending on a few factors, including the individual's pain threshold, the reason for the procedure, and the amount of sedation used during the procedure. During an EUS procedure, a thin, flexible tube with an ultrasound probe on the end is inserted into the mouth and passed down through the esophagus and stomach to the small intestine. The ultrasound probe uses high-frequency sound waves to create images of the digestive tract and surrounding tissues. Most patients are given a sedative to help them relax and feel more comfortable during the procedure. Some patients may feel mild discomfort or pressure as the scope is passed through the throat and down into the stomach, but this usually only lasts a few seconds. If any biopsies or other procedures are performed during the EUS, there may be some mild discomfort or cramping as the instruments are passed through the scope. However, most patients do not report significant pain during or after the procedure. It's important to discuss any concerns or questions you have about the EUS procedure with your healthcare provider before the procedure. They can provide more detailed information about what to expect during and after the procedure, as well as answer any specific questions you may have. | Similar | 1 | 2 | Correct but inadequate |
| How long does an EUS procedure take? | The duration of an Endoscopic Ultrasound (EUS) procedure can vary depending on several factors, including the purpose of the examination and the patient's medical condition. On average, an EUS procedure takes around Mixed with correct and incorrect/outdated data0 to 90 minutes to complete. During the EUS procedure, the patient is typically sedated to minimize discomfort. The doctor will insert an endoscope with an ultrasound probe attached through the mouth or rectum and into the gastrointestinal tract to obtain images of the organs and tissues in the area. The length of the procedure can also depend on whether any biopsies or other procedures, such as drainage of cysts, are performed during the examination. The doctor may need to spend more time collecting samples or performing additional interventions, which can extend the duration of the procedure. It's important to note that the actual duration of the EUS procedure may vary from patient to patient, so it's best to consult with your doctor to get a better idea of what to expect. | Similar | 2 | 1 | Comprehensive |
| What are the possible risks and complications of an EUS procedure? | Endoscopic ultrasound (EUS) is a minimally invasive procedure used to visualize and evaluate internal organs and structures using high-frequency sound waves. As with any medical procedure, there are potential risks and complications associated with EUS. Some possible risks and complications of EUS include: Bleeding: EUS involves the insertion of a flexible tube with an ultrasound probe attached through the mouth or rectum, which can cause small tears or cuts in the tissue. This may result in bleeding, which is usually minor and stops on its own, but in rare cases, it may require further medical intervention. Infection: EUS is generally a safe procedure, but there is a small risk of infection, especially if the endoscope is passed through the rectum. To minimize the risk of infection, the endoscope is sterilized before the procedure, and antibiotics may be given before or after the procedure. Perforation: There is a small risk of perforation or puncture of the wall of the gastrointestinal tract or other organs during the procedure. This can cause serious complications and may require surgery. Adverse reactions to sedation: EUS is typically performed under sedation, which carries its own risks. Adverse reactions to sedation can include difficulty breathing, a drop in blood pressure, and abnormal heart rhythms. Pancreatitis: In rare cases, EUS can cause pancreatitis, which is inflammation of the pancreas. This can cause severe abdominal pain, nausea, and vomiting and may require hospitalization. Other rare complications: Other rare complications of EUS include aspiration (inhaling stomach contents into the lungs), allergic reactions to the sedative or contrast dye, and damage to nearby structures such as blood vessels. It is important to discuss the risks and potential complications of EUS with your healthcare provider before the procedure. | Similar | 1 | 2 | Correct but inadequate |
| What are the alternatives to an EUS procedure? | Endoscopic ultrasound (EUS) is a medical procedure that combines endoscopy and ultrasound to examine the internal organs of the digestive tract, such as the esophagus, stomach, and pancreas. The alternative procedures to EUS depend on the specific diagnostic or therapeutic goal. Here are some common alternatives: CT scan: A computed tomography (CT) scan is a non-invasive diagnostic procedure that uses X-rays to create detailed images of the internal organs. It can be used to diagnose a wide range of conditions, including tumors, inflammation, and other abnormalities in the digestive tract. MRI: Magnetic resonance imaging (MRI) is a non-invasive diagnostic procedure that uses powerful magnets and radio waves to create detailed images of the internal organs. It can be used to diagnose a wide range of conditions, including tumors, inflammation, and other abnormalities in the digestive tract. Upper endoscopy: An upper endoscopy is a diagnostic procedure that uses a flexible tube with a camera on the end to examine the upper digestive tract, including the esophagus, stomach, and small intestine. It can be used to diagnose conditions such as ulcers, inflammation, and tumors. Colonoscopy: A colonoscopy is a diagnostic procedure that uses a flexible tube with a camera on the end to examine the colon and rectum. It can be used to diagnose conditions such as colon cancer, inflammatory bowel disease, and other abnormalities. Laparoscopy: Laparoscopy is a minimally invasive surgical procedure that uses a small camera and specialized tools to examine and treat conditions in the abdomen and pelvis. It can be used to diagnose and treat conditions such as tumors, cysts, and other abnormalities. Biopsy: A biopsy is a procedure in which a small sample of tissue is removed from the body for examination under a microscope. It can be used to diagnose conditions such as cancer, inflammation, and other abnormalities. The choice of alternative procedures depends on the specific diagnostic or therapeutic goal and the patient's individual needs and medical history. Your doctor can help you determine which procedure is best for you. | Similar | 2 | 1 | Comprehensive |
| **Treatment** |  |  |  |  |  |
| How should I prepare for my EUS procedure? | Before your EUS (Endoscopic Ultrasound) procedure, it's essential to follow your doctor's instructions carefully to ensure that the examination is as safe and effective as possible. Here are some general tips that may help you prepare: Follow your doctor's instructions: Make sure to carefully follow your doctor's instructions regarding diet, medications, and other preparations leading up to the procedure. Fasting: Generally, you will be required to fast for at least 6 hours before the procedure. This means no food or drink during this time. Medications: Inform your doctor about all the medications you are taking, including over-the-counter medications, supplements, and herbal remedies. Your doctor may ask you to stop taking certain medications temporarily before the procedure. Anesthesia: EUS is usually performed under sedation or anesthesia, which means you will need to arrange for someone to drive you home after the procedure. Wear comfortable clothing: Wear comfortable, loose-fitting clothing to the hospital or clinic on the day of your procedure. Plan for recovery time: After the procedure, you may need some time to recover from the sedation or anesthesia. Plan to take it easy for the rest of the day. Ask questions: If you have any questions or concerns about the procedure, don't hesitate to ask your doctor or the staff at the hospital or clinic. Remember to follow your doctor's instructions closely to ensure the best possible outcome for your EUS procedure. | Similar | 1 | 2 | Comprehensive |
| Can I eat or drink before an EUS procedure? | It is generally recommended that you do not eat or drink anything for at least 6 hours before undergoing an EUS (Endoscopic Ultrasound) procedure. This is to ensure that your stomach is empty, which helps to minimize the risk of complications during the procedure. If you have been instructed by your healthcare provider to take any medication before the procedure, you should follow their instructions carefully. In some cases, you may be allowed to take medication with a small sip of water. It's important to follow all instructions provided by your healthcare provider regarding fasting and preparation for the EUS procedure. If you have any questions or concerns about the procedure or its preparation, be sure to discuss them with your healthcare provider in advance. | Similar | 1 | 1 | Comprehensive |
| Will I be sedated during the EUS procedure? | During an EUS (Endoscopic Ultrasound) procedure, sedation may or may not be used, depending on the patient's preference and the complexity of the procedure. Generally, patients are given sedatives to help them relax and reduce discomfort during the procedure. Sedation may range from mild to moderate, depending on the patient's needs and the physician's discretion. The sedative is usually administered through an IV, and the patient will be monitored closely throughout the procedure to ensure their safety. However, some patients may prefer to forgo sedation and undergo the procedure while fully awake. In such cases, the physician may use a local anesthetic to numb the area being examined, which can help reduce any discomfort or pain. Ultimately, the decision to use sedation during an EUS procedure is dependent on the patient's preferences and the physician's recommendation based on the complexity of the procedure. It's important to discuss your preferences and any concerns with your physician before the procedure to ensure that you receive the most appropriate care. | Similar | 2 | 2 | Correct but inadequate |
| How long will I need to stay in the hospital after an EUS procedure? | The length of your hospital stay after an endoscopic ultrasound (EUS) procedure will depend on various factors, including the reason for the procedure, your overall health, and any potential complications that may arise during or after the procedure. In most cases, EUS is an outpatient procedure, which means you can go home the same day. However, if your doctor finds any abnormalities during the procedure, such as a suspicious lesion or mass, they may want to perform a biopsy or remove it. In this case, you may need to stay in the hospital for a day or two for observation and further treatment. It's important to follow your doctor's instructions regarding post-procedure care and activity restrictions to ensure a safe and speedy recovery. Your doctor will also provide you with specific instructions on when to resume your normal activities, including work, exercise, and diet. | Similar | 1 | 2 | Comprehensive |
| What to do for aftercare following an EUS procedure? | Endoscopic ultrasound (EUS) is a minimally invasive procedure used to examine the digestive tract, as well as other internal organs such as the pancreas and bile ducts. After the procedure, there are some aftercare instructions that patients should follow to ensure a smooth recovery. Here are some general aftercare instructions: Follow the instructions of your healthcare provider: After the procedure, your healthcare provider will give you specific instructions on how to care for yourself. Follow their advice carefully, including how to take any prescribed medications. Rest and take it easy: You may feel tired and groggy after the procedure, so it's important to rest and take it easy for a day or two. Avoid any strenuous activities and try to relax as much as possible. Monitor for any complications: In rare cases, complications can occur after an EUS procedure. Be sure to watch out for any signs of complications, such as fever, severe pain, or bleeding, and contact your healthcare provider right away if you experience any of these symptoms. Stay hydrated: Drink plenty of fluids after the procedure to help flush out any remaining sedative medication in your system. Eat a light meal: After the procedure, you should eat a light meal to help your body recover. Avoid spicy or heavy foods that may irritate your digestive tract. Arrange for transportation: You will need someone to drive you home after the procedure, as the sedative medication used during the procedure can impair your ability to drive. Follow up with your healthcare provider: Be sure to schedule a follow-up appointment with your healthcare provider to discuss the results of the procedure and any further treatment options if necessary. It's important to follow these aftercare instructions to ensure a smooth recovery and minimize the risk of complications. If you have any questions or concerns, don't hesitate to contact your healthcare provider. | Similar | 1 | 1 | Comprehensive |
| How soon can I return to my normal activities after an EUS procedure? | The recovery time after an Endoscopic Ultrasound (EUS) procedure can vary depending on the individual's health and the nature of the procedure. In general, most people can resume their normal activities within a day or two after the procedure. However, it is essential to follow the instructions given by your healthcare provider to ensure a smooth and speedy recovery. Your doctor may recommend avoiding certain foods and activities for a few days after the procedure to allow your body to heal. It is also essential to follow up with your healthcare provider after the procedure to monitor your progress and address any concerns or complications that may arise. In summary, it is best to follow the specific instructions provided by your healthcare provider, but most people can return to their normal activities within a day or two after an EUS procedure. | Similar | 1 | 1 | Comprehensive |
| How long after EUS will I receive the results of the procedure? | The timing of receiving results after an Endoscopic Ultrasound (EUS) can vary depending on several factors, including the reason for the procedure, the medical facility, and the individual case. In many cases, your doctor or healthcare provider will provide you with some initial results or impressions immediately after the procedure, while you are still in the recovery room. However, these initial results are not always conclusive or definitive, and your doctor may need to wait for more detailed analysis of the images or biopsy results. In general, it may take several days to a week or more to receive the full results of an EUS. This is because the images and any biopsy samples will need to be analyzed by a pathologist, who will prepare a report for your doctor. Your doctor will then review the report and discuss the results with you in detail during a follow-up appointment. It's important to note that you should not hesitate to contact your doctor or healthcare provider if you have any questions or concerns about your EUS results or the timing of receiving them. They can provide you with more specific information based on your individual case. | Similar | 1 | 1 | Comprehensive |
| How accurate are EUS findings in diagnosing medical conditions? | Endoscopic ultrasound (EUS) is a diagnostic imaging technique that uses a specialized endoscope equipped with an ultrasound transducer to produce high-resolution images of internal organs and structures in the gastrointestinal tract. EUS is often used to diagnose a variety of medical conditions, including cancer, pancreatic disease, and gastrointestinal disorders. The accuracy of EUS findings in diagnosing medical conditions can vary depending on several factors, including the skill and experience of the endoscopist performing the procedure, the quality of the ultrasound equipment used, and the specific condition being diagnosed. In general, however, studies have shown that EUS has a high accuracy rate in detecting and diagnosing a wide range of medical conditions. For example, a meta-analysis of studies examining the accuracy of EUS in diagnosing pancreatic cancer found that EUS had a sensitivity (the ability to detect true positive cases) of 85% and a specificity (the ability to detect true negative cases) of 95%. Another study found that EUS had a sensitivity of 90% and a specificity of 96% in diagnosing esophageal cancer. Overall, EUS is considered to be a highly accurate diagnostic tool for many medical conditions, and it is often used in conjunction with other diagnostic tests to help clinicians make an accurate diagnosis and develop an appropriate treatment plan for their patients. | Similar | 1 | 2 | Comprehensive |
| Can an EUS procedure be used to treat medical conditions? | Yes, an EUS (Endoscopic Ultrasound) procedure can be used to diagnose and treat certain medical conditions. As a diagnostic tool, EUS can be used to examine the gastrointestinal tract, including the esophagus, stomach, and small intestine, as well as nearby organs such as the pancreas, liver, and gallbladder. This can help identify abnormalities such as tumors, cysts, and inflammation. As a treatment tool, EUS can be used to perform fine needle aspiration (FNA) biopsies to obtain tissue samples for analysis, and to drain fluid collections such as cysts or abscesses. EUS can also be used to place stents to relieve blockages in the bile ducts or the gastrointestinal tract. EUS is a minimally invasive procedure that is performed using an endoscope with an ultrasound probe attached to its tip. The endoscope is passed through the mouth and into the digestive tract, allowing the physician to visualize the area of interest and perform the necessary diagnostic or therapeutic procedures. | Similar | 1 | 2 | Comprehensive |
| **Lifestyle** |  |  |  |  |  |
| Can I continue to take my medications before and after an EUS procedure? | It is generally recommended that you continue to take your medications before and after an Endoscopic Ultrasound (EUS) procedure, unless otherwise instructed by your doctor. However, it is important to inform your doctor about all the medications you are taking, including prescription medications, over-the-counter medications, and supplements. Your doctor may advise you to stop taking certain medications before the procedure, particularly if they can increase the risk of bleeding or interfere with the results of the procedure. Your doctor may also advise you to modify the timing of your medication doses before the procedure. For example, if you are taking medications that need to be taken with food, your doctor may advise you to take them earlier than usual to ensure that your stomach is empty before the procedure. After the procedure, your doctor will give you instructions on when you can resume taking your medications. In some cases, you may need to avoid certain medications for a period of time after the procedure to allow your body to recover fully. Follow your doctor's instructions carefully to ensure that you get the most benefit from your medications while minimizing any potential risks or complications. | Similar | 1 | 1 | Comprehensive |
| Are there any activities I should avoid after an EUS procedure? | After an EUS (endoscopic ultrasound) procedure, there are some activities that you should avoid to ensure a safe and smooth recovery. Here are a few things to keep in mind: Driving: You should avoid driving for at least 2Completely incorrect hours after the procedure, as the sedative used during the procedure can affect your reaction time and judgment. Operating heavy machinery: It is best to avoid operating heavy machinery or performing any tasks that require concentration and coordination until the effects of the sedative have worn off. Eating and drinking: Depending on the type of EUS procedure you had, your doctor may recommend that you avoid eating and drinking for a certain period of time after the procedure to allow your body to recover. Strenuous exercise: You should avoid any strenuous exercise or heavy lifting for at least 2Completely incorrect hours after the procedure to reduce the risk of bleeding or other complications. Drinking alcohol: It is best to avoid drinking alcohol for at least 2Completely incorrect hours after the procedure, as alcohol can interact with the sedative used during the procedure. It's always best to follow your doctor's instructions on what activities to avoid after an EUS procedure, as the specific recommendations may vary depending on your individual cas | Similar | 1 | 1 | Comprehensive |
| How can I manage pain or discomfort after an EUS procedure? | Endoscopic ultrasound (EUS) is a minimally invasive procedure that involves inserting a flexible tube with an ultrasound probe into the digestive tract. It is typically used to evaluate and diagnose conditions of the pancreas, bile duct, and gastrointestinal tract. After the procedure, some patients may experience pain or discomfort. Here are some ways to manage pain or discomfort after an EUS procedure: Take pain relievers: Over-the-counter pain relievers such as acetaminophen (Tylenol) or ibuprofen (Advil) can help relieve mild to moderate pain. If your pain is severe, your doctor may prescribe a stronger pain reliever. Apply heat: Placing a heating pad or warm towel on the area of discomfort can help ease pain and promote relaxation. Rest: Resting and avoiding strenuous activity for the first 2Completely incorrect hours after the procedure can help minimize pain and discomfort. Stay hydrated: Drinking plenty of fluids can help flush out any remaining sedatives or anesthesia and prevent dehydration, which can cause headaches and other discomfort. Follow your doctor's instructions: Your doctor may provide specific instructions on how to manage pain or discomfort after the procedure. Be sure to follow these instructions closely to ensure proper healing and recovery. If your pain or discomfort persists or worsens, or if you experience other symptoms such as fever or bleeding, be sure to contact your doctor immediately. | Similar | 1 | 2 | Correct but inadequate |
| Is it safe to travel after an EUS procedure? | Endoscopic ultrasound (EUS) is a minimally invasive diagnostic procedure that uses high-frequency sound waves to create images of the internal organs and structures in the body. The safety of traveling after an EUS procedure depends on several factors, including the type of sedation used, the complexity of the procedure, and the individual's overall health status. In general, EUS is a safe procedure with a low risk of complications. However, if sedation was used during the procedure, you may need to wait for a period of time before traveling. This is because the sedative medication used during the procedure can cause drowsiness, dizziness, and impaired coordination, which can increase the risk of falls or accidents while traveling. Your doctor will typically provide instructions on when it is safe to resume normal activities after an EUS procedure, including travel. It is important to follow these instructions carefully to avoid any potential complications. If you have any concerns about traveling after an EUS procedure, it is best to discuss them with your doctor. | Similar | 1 | 1 | Comprehensive |
| **Other** |  |  |  |  |  |
| How much does an EUS procedure cost, and will my insurance cover it? | The cost of an Endoscopic Ultrasound (EUS) procedure can vary widely depending on several factors, including the location, the facility where it is performed, and the individual physician performing the procedure. Additionally, the specific reason for the procedure, such as whether it is being done for diagnostic or therapeutic purposes, may also affect the cost. As an AI language model, I don't have access to current pricing information for specific facilities, but you can contact your healthcare provider or insurance company to get an estimate of the cost of an EUS procedure in your area. They should also be able to provide you with information on whether your insurance policy will cover the cost of the procedure. It's worth noting that insurance coverage for medical procedures can vary depending on the policy and the specific circumstances surrounding the procedure. Therefore, it's important to check with your insurance company to understand your policy's coverage and any potential out-of-pocket costs you may incur. | The cost of an EUS procedure (Endoscopic Ultrasound) can vary widely depending on various factors such as the geographical location, the facility, the specific type of EUS procedure being performed, and the individual case. In the United States, the cost of an EUS procedure can range from $2,000 to $10,000 or more. However, the actual cost can be significantly lower or higher depending on various factors such as the extent of the examination, the anesthesia used, and any additional testing or procedures that may be required. As for insurance coverage, it depends on your specific insurance plan and the reason for the EUS procedure. Generally, most insurance plans cover EUS procedures if they are deemed medically necessary. However, it is important to check with your insurance provider to confirm whether your plan covers the procedure and what your out-of-pocket expenses may be. You may also need to get pre-authorization from your insurance provider before the procedure is performed to ensure coverage. | 1 | 1 | Comprehensive |
| Can I have an EUS procedure if I am pregnant or breastfeeding? | Endoscopic ultrasound (EUS) is a medical procedure that involves the insertion of an endoscope into the gastrointestinal tract to examine organs such as the pancreas, liver, and bile ducts. If you are pregnant or breastfeeding, it is generally not recommended to undergo an EUS procedure unless it is absolutely necessary. EUS is considered a minimally invasive procedure, but there are still risks involved, including bleeding, infection, and perforation. These risks may be higher in pregnant women due to changes in the body during pregnancy. If you require an EUS procedure, it is important to discuss the potential risks and benefits with your healthcare provider. They can help determine whether the procedure is safe for you and your baby, and may recommend postponing the procedure until after pregnancy or breastfeeding has ended. | Similar | 2 | 2 | Correct but inadequate |
| What type of doctor performs an EUS procedure? | An EUS procedure, which stands for Endoscopic Ultrasound, is usually performed by a gastroenterologist who has specialized training in endoscopic procedures. Specifically, a gastroenterologist who has undergone additional training in advanced endoscopy, which includes EUS, is qualified to perform this procedure. Additionally, in some cases, a surgeon who has been trained in EUS may also perform the procedure. | Similar | 1 | 2 | Correct but inadequate |
| How can I find a qualified EUS specialist near me? | To find a qualified EUS (Endoscopic Ultrasound) specialist near you, you can try the following: Ask your primary care physician or gastroenterologist for a referral to an EUS specialist. They may have colleagues or acquaintances who specialize in this field and can recommend someone in your area. Check with local hospitals and medical centers to see if they have an EUS program or clinic. They may be able to refer you to a specialist or provide you with a list of providers in your area. Use online directories such as Healthgrades or Zocdoc to search for EUS specialists in your area. These directories usually have information on the provider's education, training, experience, and patient reviews. Consult professional organizations such as the American Society for Gastrointestinal Endoscopy (ASGE) or the American Gastroenterological Association (AGA) to find a list of EUS specialists in your area. Consider reaching out to patient support groups or online forums for advice and recommendations from other patients who have undergone EUS procedures in your area. Remember to check with your insurance provider to ensure that the specialist you choose is covered by your plan before making an appointment. | Similar | 1 | 1 | Comprehensive |
| Can I undergo an EUS procedure if I have a pacemaker or other implanted medical devices? | Electrocautery and other devices used during an endoscopic ultrasound (EUS) procedure can potentially interfere with pacemakers and other implanted medical devices. However, in most cases, undergoing an EUS procedure is considered safe for patients with pacemakers and other implanted medical devices. Prior to the procedure, your doctor will need to know about any implanted devices you have and will evaluate your individual case to determine if any precautions or modifications need to be made. Your doctor may consult with your cardiologist or other specialists to help determine if it is safe for you to undergo the procedure. During the EUS procedure, your vital signs and cardiac rhythm will be continuously monitored. Additionally, the electrosurgical equipment used during the procedure will be modified to minimize the risk of interference with your implanted device. In summary, it is generally safe for patients with pacemakers and other implanted medical devices to undergo an EUS procedure, but your doctor will need to evaluate your individual case and take appropriate precautions to ensure your safety. | Similar | 1 | 1 | Comprehensive |
| Can an EUS procedure be repeated if necessary? | Yes, an EUS (endoscopic ultrasound) procedure can be repeated if necessary. EUS is a minimally invasive diagnostic procedure that uses an endoscope equipped with an ultrasound probe to produce high-resolution images of internal organs and tissues. Repeat EUS procedures may be necessary if the initial results were inconclusive or if further evaluation or monitoring is required. The need for a repeat EUS will depend on the patient's medical condition and the judgment of the treating physician. It's worth noting that EUS is a safe and relatively low-risk procedure, but as with any medical procedure, there are some potential risks and complications. The risks associated with EUS include bleeding, infection, and perforation of the gastrointestinal tract. If you have any concerns or questions about a repeat EUS procedure, you should discuss them with your physician. | Similar | 1 | 1 | Comprehensive |
| Are there any long-term side effects or complications associated with EUS procedures? | Endoscopic ultrasound (EUS) is generally considered a safe and minimally invasive diagnostic and therapeutic procedure. However, as with any medical procedure, there are potential risks and complications that can occur. Some of the possible immediate or short-term side effects of EUS may include temporary sore throat, bloating, mild abdominal pain, nausea or vomiting, and temporary changes in bowel movements. These side effects are usually mild and go away on their own within a few hours or days. In rare cases, more serious complications may occur, such as bleeding, infection, pancreatitis, perforation of the gastrointestinal tract, or adverse reactions to sedation or anesthesia. The risk of these complications is generally low and can be minimized by choosing an experienced and skilled endoscopist, carefully following pre-procedure instructions, and closely monitoring patients during and after the procedure. As for long-term side effects or complications associated with EUS, there is currently no evidence to suggest that the procedure has any significant long-term effects. However, it is important for patients to follow post-procedure instructions and attend any scheduled follow-up appointments to ensure proper healing and monitoring. | Similar | 1 | 2 | Correct but inadequate |

Supplemental Table 4. Grading of the responses from the ChatGPT Language Model for Endoscopic retrograde cholangiopancreatography (ERCP)-related questions

| **Basic Knowledge** | 1^st^ response | 2^nd^ response | Reviewer 1 grades 1. Comprehensive 2. Correct but inadequate 3. Mixed with correct and incorrect/outdated data 4. Completely incorrect | Reviewer 1 grades 1. Comprehensive 2. Correct but inadequate 3. Mixed with correct and incorrect/outdated data 4. Completely incorrect | Reviewer 3 grades for the responses with discrepancies |
| --- | --- | --- | --- | --- | --- |
| What is ERCP | ERCP stands for Endoscopic Retrograde Cholangiopancreatography, a diagnostic and therapeutic procedure that uses an endoscope and X-rays to visualize the bile and pancreatic ducts. ERCP is done for various reasons, such as: Diagnosing and treating conditions related to the bile ducts or pancreas, such as gallstones, tumors, strictures, infections, or pancreatitis. Removing gallstones or other blockages from the bile ducts or pancreas. Obtaining biopsy samples from the bile ducts or pancreas to check for cancer or other diseases. Placing stents or other devices to relieve blockages and improve the flow of bile or pancreatic juice. ERCP is performed by a gastroenterologist who inserts an endoscope (a thin, flexible tube with a camera and a light) through the mouth and into the duodenum (the first part of the small intestine). Then, the doctor injects a dye into the bile or pancreatic ducts to make them visible on X-rays, and uses specialized instruments to perform the necessary procedures. ERCP is usually done under sedation, and most people can go home the same day. | Similar | 3 | 1 | Comprehensive |
| How is ERCP performed? | ERCP (Endoscopic Retrograde Cholangiopancreatography) is a medical procedure used to diagnose and treat conditions of the liver, gallbladder, pancreas, and bile ducts. It is performed by a gastroenterologist, who is a specialist in digestive system diseases. Here are the steps of how ERCP is performed: Pre-procedure preparation: You will be asked to fast for at least 6 hours before the procedure to avoid any complications. Your doctor may also instruct you to stop taking certain medications before the procedure. Sedation: You will be given a sedative to help you relax and reduce any discomfort during the procedure. You will be asked to lie on your left side on an exam table. Endoscopy: The doctor will insert an endoscope, a long, flexible tube with a camera at the end, through your mouth and down your esophagus to your stomach and duodenum. The camera will allow the doctor to see the inside of your digestive system and locate the opening of the bile ducts. Contrast medium injection: Once the opening of the bile ducts is located, the doctor will inject a contrast medium (dye) through the endoscope into the bile ducts. This will help to highlight any abnormalities in the bile ducts on X-rays. Imaging: The doctor will then take X-rays to visualize the contrast medium in the bile ducts. This will help to diagnose any blockages, stones, or other abnormalities in the bile ducts. Therapeutic procedures: If any abnormalities are found, the doctor may perform therapeutic procedures during the same procedure. These may include removing stones or placing a stent to keep the bile ducts open. Post-procedure care: After the procedure, you will be monitored in a recovery area until the sedative wears off. You will need to have someone drive you home as you may feel drowsy or weak for several hours after the procedure. You may also experience some mild discomfort or bloating, but these symptoms should subside within a few hours. It is important to follow your doctor's instructions carefully after the procedure to ensure proper healing and avoid any complications. | Similar | 2 | 3 | Correct but inadequate |
| How often is ERCP performed, and by whom? | ERCP (Endoscopic Retrograde Cholangiopancreatography) is a medical procedure that combines endoscopy and X-ray imaging to diagnose and treat problems related to the bile ducts, pancreas, and gallbladder. ERCP is generally performed by a gastroenterologist or a gastrointestinal (GI) surgeon who has received specialized training in this procedure. In some cases, radiologists may also perform ERCP. The frequency of ERCP procedures varies depending on the patient population and the underlying medical conditions. It is commonly used to treat conditions such as gallstones, pancreatitis, and bile duct blockages. Overall, ERCP is a relatively common procedure, with thousands of ERCPs performed each year worldwide. However, the frequency of ERCPs may vary depending on the size of the hospital or medical center, as well as the patient population they serve. | Similar | 3 | 1 | Comprehensive |
| What alternative procedures are available instead of ERCP? | Endoscopic retrograde cholangiopancreatography (ERCP) is a medical procedure used to diagnose and treat problems in the bile ducts and pancreatic ducts. However, there are several alternative procedures available, depending on the specific condition being treated. Some of these alternative procedures include: Magnetic resonance cholangiopancreatography (MRCP): This is a non-invasive imaging technique that uses MRI to visualize the bile ducts and pancreatic ducts. It can be used to diagnose problems such as gallstones, bile duct obstructions, and pancreatitis. Endoscopic ultrasound (EUS): This procedure involves using an ultrasound probe attached to an endoscope to produce detailed images of the digestive tract and surrounding tissues. It can be used to diagnose and treat conditions such as pancreatic cysts, tumors, and gallstones. Percutaneous transhepatic cholangiography (PTC): This is a minimally invasive procedure that involves inserting a needle through the skin and into the liver to inject contrast material into the bile ducts. X-rays are then used to produce images of the ducts, which can be used to diagnose and treat conditions such as bile duct obstructions. Laparoscopic cholecystectomy: This is a minimally invasive surgical procedure used to remove the gallbladder. It is typically used to treat gallstones and other conditions that affect the gallbladder. Sphincterotomy: This is a procedure that involves cutting the muscle that controls the flow of bile and pancreatic juice into the intestine. It can be used to treat conditions such as gallstones and pancreatitis. It is important to discuss the pros and cons of each procedure with your healthcare provider to determine the most appropriate course of treatment for your specific condition. | Similar | 3 | 3 | Mixed with correct and incorrect/outdated data |
| How effective is ERCP in diagnosing and treating conditions? | ERCP (Endoscopic Retrograde Cholangiopancreatography) is a medical procedure that uses an endoscope and X-rays to diagnose and treat conditions related to the bile ducts and pancreas. As a diagnostic tool, ERCP is highly effective in identifying conditions such as bile duct stones, tumors, strictures, and pancreatitis. It allows the physician to visualize the internal structures of the bile ducts and pancreas, obtain tissue samples for analysis, and take biopsies if needed. As a therapeutic tool, ERCP can be used to remove gallstones or other obstructions, dilate strictures, drain bile ducts, and place stents. It can also be used to treat pancreatitis by removing pancreatic duct stones or placing stents to relieve obstruction. Overall, ERCP is a highly effective tool for both diagnosis and treatment of conditions related to the bile ducts and pancreas. However, as with any medical procedure, there are risks associated with ERCP, such as bleeding, infection, and perforation of the digestive tract. It is important to discuss the risks and benefits of the procedure with your healthcare provider before deciding to undergo ERCP. | ERCP, which stands for Endoscopic Retrograde Cholangiopancreatography, is a diagnostic and therapeutic procedure used to examine the bile ducts, pancreas, and gallbladder. The procedure involves the use of an endoscope, which is a flexible tube with a camera attached to it, to view the inside of these organs and ducts. ERCP can be highly effective in diagnosing and treating a variety of conditions. Some of the conditions that can be diagnosed and/or treated with ERCP include: Gallstones: ERCP can be used to remove gallstones from the bile ducts. Pancreatitis: ERCP can be used to identify the cause of pancreatitis and to treat it by removing blockages or performing other procedures. Bile duct strictures: ERCP can be used to identify and treat strictures (narrowing) of the bile ducts. Bile duct and pancreatic duct leaks: ERCP can be used to identify and treat leaks in the bile ducts and pancreatic ducts. Cancer: ERCP can be used to diagnose and stage cancer of the pancreas, bile ducts, or gallbladder. Sphincter of Oddi dysfunction: ERCP can be used to diagnose and treat dysfunction of the sphincter of Oddi, a muscle that controls the flow of bile and pancreatic juice into the small intestine. Overall, ERCP can be a highly effective tool for diagnosing and treating a variety of conditions related to the bile ducts, pancreas, and gallbladder. However, like any medical procedure, it does carry some risks, and it is important to discuss these risks and potential benefits with your healthcare provider. | 3 | 2 | Correct but inadequate |
| How long does ERCP take to complete? | ERCP, or endoscopic retrograde cholangiopancreatography, is a procedure used to diagnose and treat conditions affecting the bile ducts and pancreas. The duration of an ERCP procedure can vary depending on several factors, including the complexity of the case, the experience of the medical team performing the procedure, and the patient's individual anatomy. On average, an ERCP procedure can take between 30 minutes to 2 hours to complete. However, in some cases, the procedure may take longer, especially if it involves more complex interventions such as the removal of a gallstone or the placement of a stent. Your healthcare provider can provide more specific information about how long your ERCP procedure is expected to take based on your individual case. | Similar | 1 | 1 | Comprehensive |
| What are the risks and complications of ERCP? | Endoscopic retrograde cholangiopancreatography (ERCP) is a medical procedure that is commonly used to diagnose and treat conditions affecting the bile ducts and pancreas. Like any medical procedure, ERCP comes with some risks and potential complications. Some of the risks associated with ERCP include: Pancreatitis: Pancreatitis is the most common complication of ERCP, occurring in up to 5% of cases. This is a potentially serious condition that can cause inflammation of the pancreas, leading to abdominal pain, nausea, vomiting, and fever. Bleeding: There is a small risk of bleeding during or after an ERCP procedure, particularly if a biopsy or other intervention is performed. Infection: ERCP can increase the risk of infection, particularly if the bile ducts are obstructed or there is a pre-existing infection. Perforation: In rare cases, ERCP can cause a tear or perforation in the gastrointestinal tract, which can lead to severe pain, fever, and other complications. Reaction to sedation: Some patients may experience a reaction to the sedative medication used during ERCP, which can cause respiratory depression or other complications. False-negative results: In some cases, ERCP may not be able to detect or diagnose certain conditions, leading to false-negative results. It's important to discuss the potential risks and benefits of ERCP with your doctor before the procedure, and to report any symptoms or complications promptly following the procedure. | Similar | 1 | 2 | Comprehensive |
| What happens after the ERCP procedure? | After an ERCP (Endoscopic Retrograde Cholangiopancreatography) procedure, patients are typically monitored in a recovery area for a period of time until the effects of the sedation wear off. The patient may experience some discomfort, bloating, or cramping in the abdomen after the procedure, which should gradually subside over the course of a few hours. They may also experience a sore throat or hoarseness from the tube that was inserted through their mouth. In some cases, the doctor may provide medication to help manage any discomfort or nausea. It's important for the patient to follow the post-procedure instructions provided by their doctor, which may include dietary restrictions and activity limitations for a certain period of time. The doctor will also discuss the results of the procedure with the patient, which may involve further testing or treatment depending on the findings. The patient will typically have a follow-up appointment with their doctor to discuss the results and any next steps. | Similar | 1 | 2 | Comprehensive |
| Will I need to stay in the hospital after an ERCP? | It depends on several factors, including the reason for the ERCP, your overall health, and the specific details of the procedure. In many cases, patients are able to go home the same day as the ERCP. However, some patients may need to stay in the hospital overnight for observation, especially if they have certain medical conditions or complications arise during the procedure. Your healthcare provider will be able to give you a better idea of what to expect based on your individual circumstances. They will also provide you with specific instructions on how to care for yourself after the ERCP and when to seek medical attention if necessary. | Similar | 1 | 1 | Comprehensive |
| What should I do if I experience any complications or side effects after an ERCP? | If you experience any complications or side effects after an ERCP (Endoscopic Retrograde Cholangiopancreatography), it is important to seek medical attention right away. Some of the potential complications and side effects that can occur after an ERCP include: Pancreatitis (inflammation of the pancreas) Infection Bleeding Perforation (tear or hole in the bowel) Allergic reaction to medications or contrast material used during the procedure Breathing difficulties or chest pain If you experience any of the following symptoms after an ERCP, you should contact your doctor immediately: Severe abdominal pain Nausea or vomiting Fever or chills Jaundice (yellowing of the skin or eyes) Trouble swallowing or breathing Chest pain or difficulty breathing Your doctor may recommend additional tests or treatments, depending on the specific complication or side effect you are experiencing. It is important to follow your doctor's instructions carefully and attend any follow-up appointments or check-ups as scheduled. | Similar | 1 | 2 | Correct but inadequate |
| How does the use of ERCP affect the overall prognosis of my condition? | ERCP (Endoscopic Retrograde Cholangiopancreatography) is a medical procedure that allows for visualization and treatment of the bile ducts and pancreas. The use of ERCP can be beneficial for certain medical conditions, but its impact on overall prognosis depends on the specific condition being treated. In some cases, ERCP can be used to diagnose and treat conditions such as bile duct stones, strictures, and tumors. By removing blockages or obstructions in the bile ducts or pancreas, ERCP can relieve symptoms such as abdominal pain, jaundice, and pancreatitis. In these cases, ERCP can improve the prognosis by resolving the underlying issue and preventing further complications. However, ERCP is not without risks. It is an invasive procedure that carries a small risk of complications, such as bleeding, infection, and pancreatitis. Additionally, ERCP may not always be effective in treating certain conditions, such as advanced pancreatic cancer. Therefore, the impact of ERCP on the overall prognosis of your condition depends on several factors, including the underlying medical condition, the severity of the condition, and the potential risks and benefits of the procedure. Your healthcare provider can discuss the potential benefits and risks of ERCP in your specific case and help you make an informed decision about whether the procedure is appropriate for you. | Similar | 1 | 1 | Comprehensive |
| What kind of follow-up care will I need after an ERCP procedure? | Following an endoscopic retrograde cholangiopancreatography (ERCP) procedure, you will require some post-procedure care and follow-up appointments to monitor your recovery and ensure that there are no complications. Here are some aspects of follow-up care that you may need after an ERCP procedure: Recovery: You will need to rest and avoid strenuous activities for the first 24 hours after the procedure. You may experience some discomfort, bloating, or nausea after the procedure, which should subside within a few days. Medications: Your doctor may prescribe medications to manage any pain, bloating, or nausea you may experience after the procedure. It is essential to follow the instructions for taking these medications carefully. Diet: After an ERCP procedure, you may need to avoid solid foods and stick to a liquid diet for a day or two to allow your digestive system to recover. Your doctor will give you specific dietary instructions based on your condition. Follow-up appointments: Your doctor will schedule follow-up appointments to monitor your progress after the ERCP procedure. They may also order additional tests, such as blood tests or imaging tests, to check for any complications. Watch for complications: It's crucial to keep an eye out for any signs of complications after the ERCP procedure, such as fever, severe abdominal pain, or persistent nausea and vomiting. If you experience any of these symptoms, contact your doctor immediately. In summary, after an ERCP procedure, you will need to take some time to rest and recover, follow dietary and medication instructions carefully, attend follow-up appointments, and watch for any complications. Following these steps can help ensure a smooth recovery and avoid any complications. | Similar | 1 | 2 | Comprehensive |
| **Treatment** |  |  |  |  |  |
| 6. Will I be sedated during the ERCP procedure? | ERCP (Endoscopic Retrograde Cholangiopancreatography) is a medical procedure that involves the insertion of an endoscope through your mouth and into your digestive tract. During the procedure, a sedative is usually administered to help you relax and minimize any discomfort. The type of sedative used depends on several factors, including your age, medical history, and the specific details of the procedure. Typically, a combination of sedatives and pain relievers will be given to you through an IV. This can include medications like midazolam (Versed), fentanyl, or propofol (Diprivan). The sedative medications used for ERCP will help you relax, reduce anxiety, and make the procedure more comfortable. However, it's important to note that you will not be completely unconscious during the procedure, and you may still be able to respond to instructions from the medical team. It's important to discuss any concerns you may have about sedation or the ERCP procedure with your doctor or healthcare provider beforehand. They can provide more information about the sedation process and help you understand what to expect during the procedure. | Whether or not you will be sedated during an ERCP procedure depends on a variety of factors, including your medical history and the preferences of your doctor. ERCP (Endoscopic retrograde cholangiopancreatography) is a procedure that combines endoscopy and X-ray imaging to examine the bile ducts, pancreas, and gallbladder. During the procedure, an endoscope is inserted through your mouth and down your throat, and a contrast dye is injected to help visualize the structures being examined. Sedation is typically used during an ERCP procedure to help you relax and minimize discomfort. Depending on the situation, you may receive conscious sedation, which involves the use of medications to make you feel sleepy and relaxed but still able to respond to verbal cues, or general anesthesia, which will make you fully unconscious. Your doctor will discuss the sedation options with you before the procedure and will work with you to determine the best approach based on your individual needs and medical history. It's important to follow any instructions provided by your doctor to prepare for the procedure, including any restrictions on eating or drinking beforehand. | 2 | 3 | Mixed with correct and incorrect/outdated data |
| Can ERCP be used to remove gallstones or tumors? | Yes, endoscopic retrograde cholangiopancreatography (ERCP) can be used to remove gallstones or tumors. ERCP is a procedure that combines endoscopy and X-rays to examine and treat problems in the liver, gallbladder, bile ducts, and pancreas. During an ERCP, an endoscope (a thin, flexible tube with a light and camera on the end) is passed through the mouth and into the small intestine. Then, a dye is injected into the bile ducts or pancreatic ducts to make them visible on X-rays. Using specialized tools passed through the endoscope, a doctor can remove gallstones, tumors, or other blockages from the bile ducts. ERCP can also be used to place stents (small tubes) in the bile ducts to keep them open and allow bile to flow freely. Additionally, it can be used to take a biopsy (tissue sample) of any suspicious areas to test for cancer or other diseases. It's important to note that ERCP is a complex procedure that carries some risks, including bleeding, infection, and pancreatitis. Therefore, it's typically only recommended for patients who have significant medical problems related to their bile ducts or pancreas. If you have questions about whether ERCP might be right for you, it's best to discuss them with your doctor. | Similar | 4 | 3 | Mixed with correct and incorrect/outdated data |
| What happens if the ERCP procedure doesn't provide a clear diagnosis? | If the ERCP (Endoscopic Retrograde Cholangiopancreatography) procedure does not provide a clear diagnosis, further diagnostic tests may be necessary to determine the cause of the patient's symptoms. In such cases, the doctor may recommend additional imaging tests, such as MRI (Magnetic Resonance Imaging), CT (Computed Tomography) scan, or ultrasound, to get a better view of the bile ducts and pancreas. Another possible option is to perform a biopsy during the ERCP procedure to obtain a tissue sample for further analysis. The biopsy can be used to check for the presence of cancer or other conditions that may not have been visible during the initial examination. In some cases, the doctor may repeat the ERCP procedure to get a better view of the bile ducts and pancreas or to perform additional interventions such as removing stones or placing stents. It's important to discuss the available options with your doctor and understand the risks and benefits of each before deciding on the next steps for your diagnosis and treatment | Similar | 1 | 2 | Correct but inadequate |
| How many ERCP procedures can be performed on a patient? | The number of ERCP procedures that can be performed on a patient would depend on the medical condition of the patient and the specific circumstances of each case. Endoscopic retrograde cholangiopancreatography (ERCP) is a procedure used to diagnose and treat problems in the liver, gallbladder, bile ducts, and pancreas. It involves inserting an endoscope through the mouth and into the small intestine, where a dye is injected into the bile ducts or pancreatic ducts, and X-rays are taken to look for any blockages or other abnormalities. In general, ERCP is considered a safe procedure with a low risk of complications. However, as with any medical procedure, there are potential risks involved, including bleeding, infection, perforation, and adverse reactions to sedation or anesthesia. The decision to perform additional ERCP procedures on a patient would depend on the individual patient's condition, the severity and location of the problem being treated, and the success of previous ERCP procedures. In some cases, multiple ERCP procedures may be necessary to fully diagnose or treat a condition, while in other cases, one procedure may be sufficient. Ultimately, the decision to perform additional ERCP procedures should be made by a medical professional in consultation with the patient, taking into account the risks and benefits of the procedure, as well as the patient's overall health and medical history. | Similar | 1 | 2 | Correct but inadequate |
| Can ERCP be used to treat chronic pancreatitis or pancreatic cancer? | ERCP (endoscopic retrograde cholangiopancreatography) is a diagnostic and therapeutic procedure that uses a flexible endoscope to examine the bile ducts, pancreatic ducts, and gallbladder. ERCP can be used to treat certain conditions of the pancreas and bile ducts, including chronic pancreatitis and pancreatic cancer. In chronic pancreatitis, ERCP can be used to treat blockages or strictures in the pancreatic ducts by placing stents or performing balloon dilatation. This can help relieve pain and improve the flow of pancreatic juices. In pancreatic cancer, ERCP can be used to relieve symptoms caused by obstruction of the bile ducts or pancreatic ducts. This is done by placing stents or performing other procedures to open up the ducts and allow bile or pancreatic juice to flow freely. ERCP can also be used to obtain tissue samples for diagnosis of pancreatic cancer. However, it is important to note that ERCP is not a primary treatment for pancreatic cancer or chronic pancreatitis. The treatment approach will depend on the specific condition and stage of the disease, and may involve a combination of surgery, radiation therapy, chemotherapy, or other treatments. It is best to consult with a medical professional to determine the most appropriate treatment plan for each individual case. | Similar | 2 | 3 | Mixed with correct and incorrect/outdated data |
| How is the risk of infection minimized during an ERCP procedure? | Endoscopic retrograde cholangiopancreatography (ERCP) is a procedure used to diagnose and treat conditions affecting the bile ducts, pancreatic duct, and gallbladder. During the ERCP procedure, there is a risk of infection, which can lead to serious complications. To minimize the risk of infection during an ERCP procedure, several steps are taken: Patient preparation: The patient is typically asked to fast for several hours before the procedure to reduce the risk of aspiration during sedation. They may also be given antibiotics before the procedure to prevent infection. Sterilization of equipment: All instruments used during the ERCP procedure must be sterilized or disinfected properly to prevent the spread of infection. The endoscope and other accessories used in the procedure are typically cleaned and disinfected using a high-level disinfectant. Use of sterile techniques: The procedure is performed using sterile techniques to minimize the risk of infection. The physician and other members of the medical team must wear sterile gloves, gowns, and masks. Antibiotic prophylaxis: Patients who are at a higher risk of infection may be given antibiotics before the procedure to reduce the risk of infection. Monitoring for signs of infection: Patients are closely monitored for signs of infection after the ERCP procedure. If signs of infection are detected, appropriate treatment is initiated promptly. By following these steps, the risk of infection during an ERCP procedure can be minimized. | Similar | 3 | 3 | Mixed with correct and incorrect/outdated data |
| Can I have an ERCP if I am pregnant? | ERCP, or endoscopic retrograde cholangiopancreatography, is a procedure used to diagnose and treat problems in the liver, bile ducts, and pancreas. It involves the use of an endoscope, which is a long, flexible tube with a light and camera at the end, to view the inside of these organs and ducts. During the procedure, dye is injected into the bile ducts to make them visible on X-rays. ERCP is generally not recommended during pregnancy, particularly during the first trimester, as there is a risk of harm to the developing fetus. The radiation used in X-rays can potentially harm the developing baby, and the sedatives and anesthesia used during the procedure can also be risky for the fetus. If you are pregnant and require an ERCP, your doctor will carefully evaluate the risks and benefits of the procedure and weigh them against the risks of not treating the condition. In some cases, alternative diagnostic tests or treatment options that are safer for the fetus may be recommended. It is important to discuss any concerns or questions you have with your doctor to ensure the best possible care for both you and your baby. | Similar | 1 | 2 | Comprehensive |
| **Lifestyle** |  |  |  |  |  |
| How do I prepare for an ERCP procedure? | An ERCP procedure (endoscopic retrograde cholangiopancreatography) is a medical procedure that combines endoscopy and X-ray to diagnose and treat problems in the bile ducts, pancreas, and liver. Here are some general guidelines to prepare for the procedure: Consult with your doctor: Before the procedure, consult with your doctor to ensure that the ERCP procedure is necessary and appropriate for your condition. Your doctor will also inform you about any specific preparations you need to make. Fasting: You will be required to fast for several hours before the procedure, usually for at least 6 hours. This means that you cannot eat or drink anything during this time. Medication: Inform your doctor about any medications you are taking, including prescription and over-the-counter medications, vitamins, and herbal supplements. You may need to adjust your medications before the procedure. Anesthesia: The procedure is performed under sedation or general anesthesia, so make sure to inform your doctor if you have any allergies or sensitivities to anesthesia. Arrangements for transportation: Arrange for someone to accompany you to the procedure and to drive you back home after the procedure, as you will not be able to drive yourself. Inform your doctor of any health issues: Inform your doctor of any underlying health issues, such as heart or lung problems, that may affect the procedure. Dress appropriately: Wear loose-fitting, comfortable clothing to the procedure. Follow the instructions: Follow all instructions given by your doctor regarding preparation for the procedure. It's important to follow these guidelines to ensure that the ERCP procedure is safe and effective. | Similar | 1 | 1 | Comprehensive |
| Can I eat or drink before an ERCP? | It is generally recommended to avoid eating or drinking anything for at least 6 hours prior to an ERCP (Endoscopic Retrograde Cholangiopancreatography) procedure. This is because during the procedure, a flexible tube with a camera and tools is inserted through the mouth and into the small intestine to examine and treat the bile and pancreatic ducts. Eating or drinking before the procedure may increase the risk of complications such as aspiration or vomiting during the procedure, which can be dangerous. However, it is important to follow the specific instructions provided by your healthcare provider as they may have individualized recommendations based on your medical history and the nature of your procedure. | Similar | 1 | 1 | Comprehensive |
| How soon can I resume normal activities after an ERCP? | After an ERCP (Endoscopic Retrograde Cholangiopancreatography), it is important to give your body enough time to recover from the procedure. The recovery time can vary depending on the individual and the extent of the procedure. Generally, you may need to stay in the hospital for a few hours to a day after the procedure to ensure there are no complications such as bleeding, pancreatitis or infection. During this time, it is important to follow any instructions from your healthcare provider, such as not eating or drinking until it is safe to do so. After being discharged, you should avoid driving or operating heavy machinery for at least 24 hours, as the sedative used during the procedure can cause drowsiness and affect your coordination. As for resuming normal activities, you should avoid any strenuous activities such as heavy lifting or exercise for a few days, or until your healthcare provider gives you the green light. You may also need to avoid certain foods or medications that could irritate the area where the procedure was performed. It is best to follow the specific instructions provided by your healthcare provider regarding when you can resume your normal activities. | After an ERCP (Endoscopic Retrograde Cholangiopancreatography), it is recommended to rest for a few hours until the sedative wears off. Your doctor will give you specific instructions on when you can resume normal activities based on your individual case. In general, you should avoid any strenuous activity, heavy lifting, or driving for at least 24 hours after the procedure. If you had any complications during the ERCP, such as bleeding or perforation, your recovery time may be longer, and you may need to stay in the hospital for observation. It's important to follow your doctor's instructions carefully to avoid any complications or delays in your recovery. If you have any concerns or questions, don't hesitate to contact your healthcare provider. | 1 | 2 | Correct but inadequate |
| Will I need to follow any special diet or restrictions after an ERCP? | After an ERCP (Endoscopic Retrograde Cholangiopancreatography), it is common to experience some mild digestive symptoms such as bloating, gas, and mild abdominal pain. However, in most cases, there are no specific dietary restrictions following an ERCP, and you can resume your regular diet as tolerated. However, your doctor may advise you to avoid eating or drinking anything for a few hours after the procedure to allow time for the sedative to wear off and to ensure that there are no complications from the procedure. They may also recommend avoiding heavy or greasy foods for the first few days following the procedure to reduce the risk of further digestive discomfort. If you have any specific concerns or medical conditions that require dietary restrictions, it is important to discuss this with your doctor before the procedure. They can advise you on any specific dietary considerations that may apply to your individual case. | Similar | 1 | 1 | Comprehensive |
| Can I drive myself home after an ERCP procedure? | ERCP (Endoscopic retrograde cholangiopancreatography) is a procedure used to diagnose and treat conditions of the liver, pancreas, and bile ducts. It involves the use of an endoscope, which is a long, flexible tube with a camera and a light at the end, to visualize these organs and perform interventions such as removing gallstones or inserting stents. After an ERCP procedure, you will be closely monitored for a period of time to ensure that you are recovering well from the sedation or anesthesia used during the procedure. This period of observation usually lasts for several hours, during which you will be advised not to drive or operate heavy machinery. Once the medical staff determines that you are stable and alert enough to go home, you will be given discharge instructions. It is generally recommended that you have a friend or family member drive you home, rather than driving yourself. This is because the sedation or anesthesia used during the procedure can impair your reflexes, coordination, and judgment, which could make it unsafe for you to drive. It's important to follow your doctor's instructions carefully and avoid driving or operating heavy machinery until you have fully recovered from the effects of the sedation or anesthesia. If you have any concerns or questions about driving after an ERCP procedure, you should discuss them with your doctor or the medical staff who are overseeing your care. | Similar | 1 | 1 | Comprehensive |
| Can I take my regular medication before and after an ERCP? | It's important to consult with your doctor or healthcare provider about whether you should take your regular medication before and after an ERCP (endoscopic retrograde cholangiopancreatography) procedure. Some medications may need to be stopped or adjusted before the procedure due to potential interactions with the medications used during ERCP, such as sedatives or pain medications. Additionally, some medications may affect the liver or bile ducts, which can be examined during the procedure. Your doctor may also advise you to stop taking certain medications after the procedure to reduce the risk of complications, such as bleeding. Therefore, it is important to discuss any concerns or questions about your medication regimen with your healthcare provider before and after an ERCP. | Similar | 1 | 1 | Comprehensive |
| **Others** |  |  |  |  |  |
| Is ERCP painful? | Endoscopic retrograde cholangiopancreatography (ERCP) is a medical procedure that is performed to diagnose and treat problems in the liver, gallbladder, bile ducts, and pancreas. ERCP can be uncomfortable, and some people may experience pain during or after the procedure, but it is usually well-tolerated with the use of sedation or anesthesia. During the procedure, the doctor will pass a flexible tube called an endoscope through your mouth and down your throat to reach the small intestine. The endoscope has a camera on the end that allows the doctor to see inside your body and guide a thin tube called a catheter into the bile ducts. The doctor will then inject a contrast dye into the catheter, which will help highlight any blockages or abnormalities on x-ray images. The use of sedation or anesthesia will help to make the procedure more comfortable and reduce any pain or discomfort you may feel during the procedure. After the procedure, you may experience mild discomfort, such as bloating or cramping, but this should subside within a few hours. In rare cases, more serious complications can occur, such as bleeding, infection, or perforation of the intestine, which can cause more significant pain. Your doctor will discuss the risks and benefits of ERCP with you before the procedure and take steps to minimize any discomfort or pain. | Similar | 1 | 2 | Comprehensive |
| Will I need someone to accompany me to and from the hospital for the ERCP procedure? | It is generally recommended to have someone accompany you to and from the hospital for any medical procedure, including the ERCP (endoscopic retrograde cholangiopancreatography) procedure. During the ERCP procedure, you will be given sedation, and the effects of the medication can last for several hours afterward. It may not be safe for you to drive or travel alone during this time, so having someone with you to help you get home safely is important. Additionally, after the ERCP procedure, you may experience side effects such as dizziness, nausea, or discomfort, and having someone with you can provide emotional support and assistance if needed. It is best to check with your healthcare provider or the hospital where the procedure is being performed to confirm their specific recommendations for accompanying someone for an ERCP procedure. | Similar | 1 | 1 | Comprehensive |
| How much does an ERCP procedure cost and will my insurance cover it? | The cost of an ERCP (Endoscopic Retrograde Cholangiopancreatography) procedure can vary depending on several factors, including the location of the medical facility, the complexity of the procedure, and the individual's insurance coverage. On average, an ERCP can cost between $2,500 and $10,000 in the United States, although prices may be higher in some areas. It's important to note that this cost does not include additional expenses such as anesthesia, follow-up appointments, and other related medical services. As for insurance coverage, it will depend on your specific policy and plan. Some insurance companies may cover the cost of an ERCP procedure entirely, while others may only cover a portion of the cost. You should contact your insurance provider directly to learn about your coverage options and any out-of-pocket costs you may be responsible for. It's also worth noting that many hospitals and medical facilities offer financial assistance programs for those who are unable to afford the full cost of a procedure. You can inquire about these programs with your medical provider or hospital billing department. | Similar | 1 | 1 | Comprehensive |

Supplemental Table 5: Grading responses to emotional support questions by ChatGPT and GPT-4 models

| EUS Procedure | ChatGPT | Grading. (Scale 1-5, 5 being most comprehensive answer, 1 being least comprehensive) | GPT-4 | Grading. (Scale 1-5, 5 being most comprehensive answer, 1 being least comprehensive) |
| --- | --- | --- | --- | --- |
| As someone with an upcoming EUS procedure, I feel quite nervous. Can you suggest ways to help me emotionally prepare and feel more at ease about the process? | Undergoing a medical procedure can be an anxious experience, and it's perfectly normal to feel nervous about it. Here are some tips to help you emotionally prepare and feel more at ease about the upcoming EUS procedure: Educate yourself: Research the procedure and its purpose. Knowing what to expect and why the procedure is necessary can help alleviate some anxiety. Talk to your doctor: Ask your doctor any questions you may have about the procedure, and what you can expect before, during, and after the procedure. They can provide you with information about the procedure and what to expect during it. Practice relaxation techniques: Deep breathing, meditation, and other relaxation techniques can help calm your nerves and ease your anxiety. Try incorporating these techniques into your daily routine leading up to the procedure. Seek support: Talk to friends, family, or a mental health professional about your fears and concerns. Having someone to talk to can help you feel more supported and less alone. Visualize a positive outcome: Visualization can be a powerful tool for reducing anxiety. Try to imagine the procedure going smoothly and visualize a positive outcome. Take care of yourself: Make sure you are eating well, getting enough sleep, and exercising regularly. Taking care of yourself can help you feel more physically and emotionally prepared for the procedure. Remember, it's normal to feel anxious before a medical procedure, but there are many things you can do to help alleviate your fears and prepare yourself emotionally for the EUS procedure. | 4 | It's completely normal to feel nervous about an upcoming endoscopic ultrasound (EUS) procedure. Here are some suggestions to help you emotionally prepare and feel more at ease about the process: Educate yourself: Understand the procedure, its benefits, and potential risks. Knowing what to expect can help alleviate some of your anxiety. Speak with your doctor and ask any questions you may have. Communicate your concerns: Share your feelings and concerns with your doctor, nurse, or other healthcare providers. They can provide reassurance, answer questions, and address any fears you may have. Seek support: Talk to friends, family, or a professional counselor about your feelings. Sometimes, sharing your concerns with others can help put things into perspective and provide emotional support. Practice relaxation techniques: Deep breathing, meditation, or progressive muscle relaxation can help you relax and manage stress. Consider practicing these techniques daily leading up to the procedure. Visualize a positive outcome: Spend some time each day visualizing the procedure going smoothly and successfully. This can help create a sense of calm and optimism. Maintain a healthy lifestyle: Get plenty of sleep, eat well, and exercise regularly. A healthy body can help support a healthy mind. Stay busy: Keep yourself occupied with activities you enjoy, such as hobbies or spending time with loved ones. This can help take your mind off the procedure and reduce anxiety. Trust your medical team: Remind yourself that the healthcare professionals performing the procedure are trained and experienced. Trust in their expertise to ensure your safety and well-being. Prepare for recovery: Understand the post-procedure instructions and make any necessary arrangements for rest and recovery. Knowing you are prepared can help you feel more at ease. Consider sedation options: If you're particularly anxious, talk to your doctor about sedation options for the procedure. This can help make the experience more comfortable and less stressful. Remember, it's normal to feel nervous before a medical procedure. By following these suggestions and keeping an open line of communication with your healthcare team, you can help reduce your anxiety and feel more prepared for your EUS procedure. | 5 |
| My spouse is scheduled for an EUS procedure, and we're feeling anxious about the diagnosis. What can we do to cope with this uncertainty and fear ? | It's understandable to feel anxious and uncertain about a medical procedure that your spouse is about to undergo. Here are some suggestions that may help you both cope with the situation: Educate yourself: Learn as much as you can about the procedure your spouse is scheduled for, including what to expect during and after the procedure. This can help reduce anxiety by giving you a sense of control and familiarity with the process. Communicate with your healthcare provider: Ask questions and voice your concerns to your healthcare provider. They can help alleviate some of your fears and provide you with more information about the procedure. Seek support: Reach out to friends and family for emotional support. Consider joining a support group or talking with a therapist to help you cope with the uncertainty and anxiety. Practice self-care: Take care of your physical and emotional health. Get enough sleep, exercise regularly, eat a healthy diet, and engage in activities that bring you joy and relaxation. Stay positive: Try to focus on positive outcomes and remain hopeful. Keep in mind that many medical procedures have successful outcomes, and your healthcare team will do everything possible to ensure the best possible outcome for your spouse. Remember, it's normal to feel anxious and uncertain about a medical procedure, but by taking proactive steps to educate yourself, seek support, and practice self-care, you can help reduce your anxiety and cope with the situation more effectively. | 4 | I understand that it's natural to feel anxious and fearful about a medical procedure and potential diagnosis. Here are some suggestions to help both of you cope with the uncertainty: Educate yourselves: Learn about the EUS (endoscopic ultrasound) procedure, why it's being done, and what the possible outcomes might be. Understanding the procedure can help alleviate some of the fear. Communicate: Talk openly about your concerns and feelings with each other. Having an honest conversation can provide support and reassurance. Seek professional support: If you or your spouse are struggling to cope with the uncertainty, consider reaching out to a mental health professional, such as a psychologist or therapist, who can provide guidance and coping strategies. Reach out to loved ones: Share your concerns with close friends or family members. They may provide emotional support, encouragement, and reassurance. Practice stress reduction techniques: Engage in activities such as meditation, yoga, or deep breathing exercises to help reduce anxiety and stress. Maintain a routine: Keeping up with daily routines can provide a sense of stability and normalcy during uncertain times. Stay active: Engage in physical activity to help relieve stress and improve your overall well-being. Focus on what you can control: While you cannot control the outcome of the EUS, you can control how you react and the steps you take to care for yourself and your spouse. Be patient: It's essential to remember that waiting for results is a normal part of the process. Try to remain patient and focus on the present moment. Plan for different outcomes: While it's natural to hope for the best, it can be helpful to consider different scenarios and how you would cope with each. This can help you feel more prepared for whatever the outcome may be. Remember, it's normal to feel anxious about a medical procedure and its potential diagnosis. Try to focus on the things you can control, and don't hesitate to seek support from loved ones or professionals if needed. | 5 |
| After receiving a concerning diagnosis from my recent EUS procedure, I am feeling scared and overwhelmed. Can you help me process my emotions? | I'm sorry to hear that you're feeling scared and overwhelmed after receiving a concerning diagnosis from your recent EUS procedure. It's completely understandable to feel this way, and it's important to take the time to process your emotions. One thing that may be helpful is to talk to someone about how you're feeling. This could be a trusted friend or family member, a therapist, or even a support group for people going through similar experiences. Sharing your feelings with someone who understands and cares about you can help you feel less alone and more supported. It's also important to take care of yourself during this time. This could mean getting enough rest, eating well, and engaging in activities that you enjoy. Exercise can also be a great way to relieve stress and improve your mood. Finally, try to stay positive and focus on what you can do to improve your health. This may involve making lifestyle changes, following your doctor's recommendations, and seeking out additional medical care or support as needed. Remember, it's normal to feel scared and overwhelmed after receiving a concerning diagnosis, but there are things you can do to cope with these emotions and take control of your health. | 3 | I'm not a mental health professional, but I'll do my best to help you process your emotions. It's completely normal to feel scared and overwhelmed after receiving a concerning diagnosis. Here are a few suggestions that may help you cope: Allow yourself to feel: Give yourself permission to experience your emotions, as they are a natural response to the situation. It's important to acknowledge and validate your feelings. Reach out for support: Talk to your friends, family, or a mental health professional about what you're going through. It can be helpful to share your feelings with people you trust and who can offer emotional support. Educate yourself: Learn more about your diagnosis so you can better understand your condition and its implications. Consult with your healthcare team to gather accurate information and avoid misinformation. Focus on what you can control: While there might be aspects of your diagnosis that are out of your control, focus on the things you can influence, such as following your healthcare provider's recommendations, maintaining a healthy lifestyle, and seeking emotional support. Set realistic goals: Break down your challenges into smaller, manageable tasks. This can help you focus on achievable goals and feel a sense of accomplishment. Practice self-care: Make time for relaxation, engage in activities that you enjoy, and maintain a healthy sleep schedule. Taking care of your physical and mental well-being is crucial during challenging times. Consider joining a support group: Connecting with others who are facing similar situations can provide a sense of community and understanding. Remember that it's okay to seek help and support from others, and it's important to address your emotions as you navigate this challenging time. If you find that your emotions are becoming too difficult to manage, consider reaching out to a mental health professional for guidance. | 5 |
| The idea of an EUS procedure terrifies me, and I am afraid of potential complications. Is there any way to manage my fear and anxiety? | It's normal to feel anxious or fearful about medical procedures, especially if you don't know much about them. However, there are several ways to manage your fear and anxiety about an EUS (Endoscopic Ultrasound) procedure. Educate yourself: Learn as much as you can about the procedure, including what it entails, how it is performed, and the potential risks and benefits. Your doctor can provide you with information, and you can also research online. Talk to your doctor: Discuss your concerns with your doctor, and ask any questions you may have. They can help address any fears or concerns you may have and provide reassurance. Practice relaxation techniques: Before and during the procedure, try deep breathing exercises, visualization, or other relaxation techniques to calm your nerves. Seek support: Talk to family members or friends who have undergone similar procedures or join a support group to connect with others who have had EUS or other medical procedures. Consider medication: If your anxiety is severe, talk to your doctor about medications that may help you relax before the procedure. Remember, EUS is a common and safe medical procedure, and most patients experience little to no complications. By taking steps to manage your fear and anxiety, you can help ensure a successful and comfortable procedure. | 4 | It's normal to feel fear and anxiety about medical procedures like endoscopic ultrasound (EUS). To help manage your fear and anxiety, you can consider the following strategies: Education: Learn about the EUS procedure, its benefits, and potential risks. Understanding the procedure can help alleviate some of your fears. Communication: Talk to your healthcare team about your concerns. They can provide information, reassurance, and answer any questions you may have. Relaxation techniques: Practice deep breathing, meditation, or progressive muscle relaxation to help reduce anxiety and stress. Visualization: Imagine yourself going through the procedure calmly and successfully. This mental rehearsal can help build confidence and ease anxiety. Support: Share your feelings with friends, family, or support groups. They can offer comfort, encouragement, and practical advice. Distraction: Engage in activities that take your mind off the procedure, such as reading, watching movies, or listening to music. Professional help: If your anxiety is severe or persistent, consider seeking help from a mental health professional, such as a psychologist or psychiatrist. Ask about sedation: Talk to your healthcare provider about sedation options during the EUS procedure, which can help you feel more relaxed and comfortable. Remember, it's essential to discuss your feelings and concerns with your healthcare provider, who can help you make the best decisions for your situation. | 5 |
| Endoscopy |  |  |  |  |
| As my endoscopy approaches, I'm feeling very anxious about the results | It's completely understandable to feel anxious before a medical procedure like an endoscopy. It's natural to worry about the results, especially if you're not sure what to expect. However, it's important to remember that the vast majority of endoscopies are routine and don't reveal any serious problems. Here are a few tips that may help ease your anxiety: Talk to your doctor: Before the procedure, make sure to discuss any concerns or questions you may have with your doctor. They can help explain what to expect and provide reassurance about the procedure. Practice relaxation techniques: Deep breathing, meditation, and other relaxation techniques can help you manage anxiety. Try to practice these techniques in the days leading up to the procedure. Lean on support: Reach out to friends and family members for support. Talking about your fears and concerns can help you feel less alone. Keep yourself distracted: Try to keep yourself occupied with activities that you enjoy. This can help take your mind off the procedure and the results. Remember that whatever the results may be, you will have the support and care of your doctor to help you navigate any necessary next steps. | 4 | It's completely normal to feel anxious about an upcoming endoscopy and its results. The anticipation and uncertainty can be stressful, but it's important to remember that the procedure is a diagnostic tool designed to help your doctor identify and treat any issues you may be experiencing. To help manage your anxiety, consider trying the following strategies: Educate yourself: Learn about the endoscopy procedure, its purpose, and what to expect. Knowing what will happen during the procedure can help alleviate some of the fear of the unknown. Ask questions: If you have concerns or questions about the procedure, don't hesitate to discuss them with your doctor. They can provide reassurance and address any misconceptions you might have. Focus on the positive: Remind yourself that the endoscopy is an important step toward understanding and potentially treating any issues you may be facing. It's a proactive measure to ensure your health and well-being. Practice relaxation techniques: Deep breathing exercises, meditation, and progressive muscle relaxation can help calm your mind and reduce anxiety. Talk to someone: Share your feelings with a trusted friend or family member. Sometimes, discussing your concerns with someone who cares about you can help put things into perspective and provide emotional support. Stay busy: Engage in activities you enjoy or find relaxing, such as reading, watching movies, or spending time with friends and family. Staying occupied can help distract you from your worries. Consider professional support: If your anxiety becomes overwhelming, consider speaking with a mental health professional, such as a therapist or counselor, to help you develop coping strategies. Remember, it's normal to feel anxious about medical procedures, but taking steps to manage your anxiety can make the experience more manageable. | 5 |
| I recently received abnormal endoscopy results and am struggling with anxiety and sadness. | I'm sorry to hear that you received abnormal endoscopy results and that you're struggling with anxiety and sadness. It's completely normal to feel this way when faced with health concerns. It's important to take care of both your physical and mental health during this time. Firstly, it's important to follow up with your doctor and discuss your endoscopy results. Your doctor can provide you with more information about your results, answer any questions you may have, and recommend any necessary treatment options. Secondly, it's important to take care of your mental health during this time. Anxiety and sadness can be overwhelming, but there are things you can do to manage these feelings. Here are some tips: Practice self-care: Make sure to prioritize your physical and mental health by getting enough sleep, eating a balanced diet, and engaging in activities that make you feel good. Stay connected: Reach out to friends and family for support. Talking about your feelings can help you feel less alone. Seek professional help: Consider talking to a mental health professional such as a therapist or counselor. They can help you develop coping strategies and provide support during this difficult time. Practice mindfulness: Engage in activities such as meditation, yoga, or deep breathing exercises to help manage anxiety. Remember, it's okay to feel anxious and sad. Take things one step at a time and remember that you are not alone. | 5 | I'm not a doctor, but I can try to help you cope with the anxiety and sadness you're experiencing due to your abnormal endoscopy results. It's completely natural to feel this way, especially when dealing with uncertainty about your health. Here are a few suggestions to help you manage your emotions: Reach out to your support network: Share your feelings with friends and family who can provide emotional support and encouragement. You don't have to go through this alone. Communicate with your doctor: Make sure you have a thorough understanding of your endoscopy results and their implications. Ask your doctor any questions you may have and discuss potential next steps. Focus on what you can control: While you can't control the outcome of the endoscopy, you can control how you take care of yourself. Focus on maintaining a healthy lifestyle, including eating well, getting regular exercise, and practicing good sleep habits. Practice relaxation techniques: Techniques such as deep breathing, progressive muscle relaxation, or meditation can help reduce anxiety and promote a sense of calm. Seek professional help if necessary: If your anxiety and sadness persist or worsen, consider talking to a mental health professional who can provide guidance and support tailored to your specific situation. Remember, it's essential to follow your doctor's advice and recommendations for any medical concerns. | 5 |
| With a history of panic attacks, I'm concerned about my upcoming endoscopy. | It's completely understandable to feel anxious about an upcoming medical procedure, especially if you have a history of panic attacks. Endoscopies are generally safe and routine procedures, but it's still important to take steps to manage your anxiety and make the experience as comfortable as possible. Here are a few tips that might help: Talk to your doctor: Let your doctor know about your history of panic attacks and your concerns about the procedure. They may be able to offer you some reassurance or prescribe medication to help you stay calm during the procedure. Practice relaxation techniques: Deep breathing, progressive muscle relaxation, and visualization are all techniques that can help you calm your mind and body. You might try practicing these techniques before the procedure to help reduce your anxiety. Bring a support person: If possible, bring a trusted friend or family member with you to the procedure. Knowing that someone is there to support you can be very comforting. Use distraction techniques: Listening to music, watching a favorite TV show, or reading a book can help distract your mind from your anxiety and make the time pass more quickly. Remember, it's important to take care of both your physical and mental health. Don't hesitate to reach out to your healthcare provider if you have any concerns or questions about the procedure. | 5 | It's completely normal to feel anxious or concerned about an upcoming medical procedure, especially if you have a history of panic attacks. Here are some tips to help you manage your anxiety and prepare for your endoscopy: Educate yourself: Learn about the endoscopy procedure, its purpose, and what to expect. Understanding the process can help alleviate some of your concerns. Speak with your doctor: Share your concerns with your doctor or the medical team performing the endoscopy. They can provide additional information, reassurance, and address any specific concerns you have. Practice relaxation techniques: Deep breathing exercises, meditation, or progressive muscle relaxation can help manage anxiety and stress leading up to the procedure. Bring a support person: Having a friend or family member with you on the day of the procedure can provide emotional support and help alleviate anxiety. Ask about sedation: Depending on the type of endoscopy you're having, sedation or anesthesia may be offered to help you feel more comfortable and relaxed during the procedure. Discuss your options with your doctor. Establish a post-procedure plan: Knowing what to expect after the endoscopy, including any necessary follow-up appointments, can help reduce anxiety. Remember, it's important to communicate your concerns with your healthcare team so they can address them and ensure you're as comfortable as possible during the procedure. | 4 |
| Colonoscopy |  |  |  |  |
| The idea of having a colonoscopy makes me feel extremely uncomfortable and anxious. | t's understandable to feel uncomfortable and anxious about the idea of having a colonoscopy. However, it's important to remember that this is a routine medical procedure that can be essential for maintaining good health. If you are feeling anxious, it may be helpful to talk to your doctor or healthcare provider about your concerns. They can provide you with more information about the procedure and help address any specific fears or questions you may have. Additionally, there are some practical steps you can take to help make the experience more comfortable. For example, you may want to ask about sedation options to help you relax during the procedure, or bring a trusted friend or family member to provide emotional support. Remember that colonoscopies are a common and important medical procedure that can help detect and prevent serious health conditions, such as colon cancer. By taking care of your health and getting the necessary screenings, you are taking an important step towards maintaining your overall well-being. | 3 | It's completely normal to feel anxious and uncomfortable about the idea of having a colonoscopy. Medical procedures can be intimidating, especially when they involve sensitive areas of the body. However, a colonoscopy is an important screening tool for detecting colorectal cancer and other conditions early, when they are most treatable. Here are some tips to help you manage your anxiety and discomfort: Educate yourself: Learn about the procedure, its benefits, and how it's performed. Understanding what to expect can help alleviate some of your fears. Speak with your doctor: Address your concerns with your healthcare provider. They can provide you with information and reassurance to help ease your anxiety. Relaxation techniques: Practicing deep breathing exercises, meditation, or progressive muscle relaxation can help reduce anxiety. Distraction: Find ways to distract yourself from your worries, such as reading, listening to music, or engaging in hobbies. Seek support: Talk to friends or family members who have had a colonoscopy about their experiences. They may be able to provide you with some insight and reassurance. Consider sedation: During a colonoscopy, you may be offered sedation to help you relax. Discuss your options with your doctor. Focus on the benefits: Remind yourself of the importance of the procedure and how it can help prevent serious health issues. Remember, it's okay to feel anxious about medical procedures. Discussing your concerns with your healthcare provider and practicing relaxation techniques can help make the experience more manageable. | 5 |
| My partner is scheduled for a colonoscopy, and we are both concerned about the potential findings. | It's understandable to feel concerned about the potential findings of a colonoscopy. However, it's important to remember that a colonoscopy is a routine procedure that is used to screen for and detect colon cancer, as well as other conditions such as polyps and inflammatory bowel disease. If any abnormalities are found during the colonoscopy, your partner's doctor will discuss the findings with you and recommend further testing or treatment if necessary. It's important to remember that early detection and treatment of colon cancer and other conditions can improve outcomes and increase the likelihood of successful treatment. In the meantime, you can help your partner prepare for the procedure by following the instructions provided by their doctor or healthcare provider, such as fasting before the procedure and taking any prescribed medications. You can also offer emotional support and be there for them throughout the process. | 3 | It's natural to be concerned about the potential findings of a colonoscopy, as it can detect various conditions, some of which might be serious. A colonoscopy is a valuable diagnostic tool that helps doctors examine the lining of the colon and rectum to identify any abnormalities, such as polyps, inflammation, or cancer. Some potential findings of a colonoscopy include: Normal colon: In many cases, the colonoscopy reveals a healthy colon without any abnormalities. Polyps: These are small growths on the lining of the colon that can be benign (noncancerous) or precancerous. If polyps are detected, they are often removed during the procedure to prevent the possibility of them developing into cancer. Inflammatory bowel disease (IBD): A colonoscopy can help diagnose conditions like Crohn's disease or ulcerative colitis, which are chronic inflammatory conditions of the digestive tract. Diverticulosis: This condition occurs when small pouches (diverticula) form in the colon's lining, which can sometimes become inflamed or infected. Colorectal cancer: A colonoscopy is an essential tool in diagnosing colorectal cancer. If any suspicious growths or tissue are found, they will be biopsied and analyzed to determine if they are cancerous. It's important to remember that many colonoscopies result in normal or benign findings. However, even if an issue is discovered, early detection is crucial for successful treatment. It's better to know and address any potential problems rather than ignoring them. If you or your partner have concerns, it's always a good idea to discuss them with the healthcare provider performing the colonoscopy. They can provide additional information and reassurance about the procedure and its potential outcomes. | 5 |
| I recently had a colonoscopy and received a worrisome diagnosis. I am feeling frightened and overwhelmed. | I'm sorry to hear that you received a worrisome diagnosis after your colonoscopy. It's completely normal to feel frightened and overwhelmed in a situation like this. It's important to remember that you are not alone and that there are many resources available to help you cope with your diagnosis. One important step you can take is to schedule an appointment with your doctor to discuss your diagnosis in more detail. It may be helpful to bring a family member or friend with you for support. You may also want to consider seeking out a support group for people with your condition. Support groups can provide a safe and supportive environment to share your feelings and experiences with others who understand what you're going through. It's also important to take care of yourself during this time. Make sure you are getting enough rest, eating a healthy diet, and engaging in activities that bring you joy and relaxation. Remember that while a diagnosis can be frightening, it is also the first step in getting the treatment you need to manage your condition. With the right care and support, many people are able to live full and healthy lives with a chronic illness. | 4 | I'm not a doctor, but I can try to help you find some information and support. First of all, it's completely normal to feel frightened and overwhelmed after receiving a worrisome diagnosis. It's important to remember that you're not alone, and there are resources available to help you cope with your emotions and manage your health. Understand your diagnosis: Educate yourself about your condition by discussing it with your doctor, reading trusted sources, or seeking information from relevant medical organizations. This will help you better understand what to expect and how to manage your health moving forward. Seek emotional support: Talk to your friends, family, or a mental health professional about your feelings. It's important to have a support system in place to help you navigate this challenging time. Join a support group: Connect with others who have been through similar experiences. Many organizations offer support groups for individuals with specific medical conditions, where you can share your feelings, ask questions, and learn from others. Focus on self-care: Make sure you are taking care of your physical and mental health. Get enough rest, eat a balanced diet, exercise, and engage in activities that bring you joy and relaxation. Stay organized: Keep track of your medical appointments, test results, and treatment plans in a journal or digital app. This will help you stay on top of your health care and make informed decisions. Be proactive: Work closely with your healthcare team to develop a treatment plan that works best for you. Ask questions and advocate for your needs. Stay hopeful: While it's natural to feel worried, try to stay positive and focus on what you can control. There are often many treatment options available, and medical advancements are continually being made. Remember that it's important to reach out to your healthcare provider for guidance and support. They will be able to give you the most accurate information and advice tailored to your specific situation. | 5 |
| My mother needs to have a colonoscopy, but she is terrified of the procedure. | I understand that your mother may be feeling scared about the colonoscopy procedure. However, it's important to remember that colonoscopies are a common and important medical procedure used to detect and prevent colon cancer. Here are some tips to help your mother feel more comfortable about the procedure: Explain the procedure in detail: It may be helpful to explain the entire process to your mother so she knows what to expect. You can describe how the procedure is done, how long it takes, and what she can expect to feel during the procedure. Talk to the doctor: Encourage your mother to talk to the doctor about any concerns she may have. The doctor can explain the procedure and answer any questions she may have. Discuss sedation options: Many colonoscopy procedures are done with sedation, which can help your mother feel more comfortable during the procedure. Talk to the doctor about what sedation options are available and which one may be best for your mother. Bring a support person: Your mother may feel more comfortable if she has a trusted person with her during the procedure. Consider going with her to the appointment or arranging for someone else to be there with her. Focus on the benefits: Remind your mother that a colonoscopy can help detect and prevent colon cancer, which is much more serious than any temporary discomfort from the procedure. Distractions: Some medical facilities may offer entertainment or other distractions to help patients during the procedure. You can ask the medical staff about any options available to help your mother feel more comfortable. Remember that it's natural to feel scared or nervous about medical procedures, but there are steps you can take to help your mother feel more comfortable and confident about the colonoscopy. | 5 | It's completely normal for your mother to be anxious about a colonoscopy, as many people feel nervous about medical procedures. Here are some suggestions to help alleviate her fears: Education: Learn about the procedure together. Understanding the process and its importance in detecting colon-related issues, such as polyps or colorectal cancer, can help reduce anxiety. Speak with the doctor: Encourage your mother to discuss her concerns with her physician. The doctor can provide information, answer questions, and help her feel more at ease about the procedure. Sedation: Most colonoscopies are performed under sedation or anesthesia, which helps patients feel more comfortable and relaxed during the procedure. Make sure your mother is aware of this, as it can help ease her fears. Support: Offer to accompany your mother to the procedure and provide emotional support. Having someone she trusts by her side can help her feel more secure. Share experiences: If you know someone who has had a colonoscopy, encourage them to share their experience with your mother. Hearing positive stories can help dispel misconceptions and provide reassurance. Preparation: Help your mother with the necessary preparations, such as following the prescribed diet and bowel preparation regimen. Feeling prepared can help reduce anxiety. Relaxation techniques: Encourage your mother to practice deep breathing exercises, visualization, or meditation to help her relax in the days leading up to the procedure. Distraction: Encourage your mother to engage in enjoyable activities that will take her mind off the upcoming procedure. Remember, it's essential to empathize with her fears and be patient as she works through her anxiety. | 5 |
| ERCP |  |  |  |  |
| I am feeling anxious about the ERCP procedure and have concerns about the procedure. | I'm sorry to hear that you're feeling anxious about the ERCP procedure. It's completely normal to feel anxious before any medical procedure, especially one that involves a certain degree of risk. To help alleviate your concerns, it may be helpful to learn more about the ERCP procedure and what it entails. ERCP stands for Endoscopic Retrograde Cholangiopancreatography, and it's a minimally invasive procedure that is commonly used to diagnose and treat conditions related to the liver, bile ducts, and pancreas. During the procedure, a flexible tube with a camera at the end (an endoscope) is inserted through your mouth and guided down your throat and into your stomach and small intestine. Once the endoscope is in place, a small tube is inserted through the endoscope and into the bile ducts or pancreatic ducts, allowing the doctor to inject contrast dye and take X-ray images. While the ERCP procedure does carry some risks, such as bleeding, infection, and pancreatitis, these risks are generally considered low. Additionally, your doctor will take all necessary precautions to minimize these risks and ensure your safety during the procedure. If you have any specific concerns or questions about the ERCP procedure, it's important to discuss them with your doctor. They can provide you with more information about the risks and benefits of the procedure, as well as address any other concerns you may have. They may also be able to offer you some tips or techniques for managing your anxiety before, during, and after the procedure. | 4 | I'm not a medical professional, but I can provide some general information about the ERCP procedure that might help you understand it better. Keep in mind that you should consult with your healthcare provider for personalized advice and recommendations. ERCP (Endoscopic Retrograde Cholangiopancreatography) is a diagnostic and treatment procedure that uses an endoscope and X-rays to examine the bile and pancreatic ducts. It's typically used to diagnose and treat conditions like gallstones, bile duct stones, chronic pancreatitis, and bile or pancreatic duct obstruction. It's normal to feel anxious about any medical procedure, especially if it's your first time. Here are some things you can do to help manage your anxiety: Educate yourself: Understanding the procedure, its benefits, and potential risks can help you feel more in control and reduce anxiety. Don't hesitate to ask your healthcare provider any questions you may have. Communicate your concerns: Talk openly with your healthcare provider about your anxiety and concerns. They can help address your questions and provide reassurance. Bring support: If possible, have a trusted friend or family member accompany you to the procedure. Their presence can provide emotional support and help you feel less anxious. Practice relaxation techniques: Deep breathing, meditation, or progressive muscle relaxation can help calm your nerves before the procedure. Focus on positive thoughts: Think about the potential benefits of the procedure, such as improved health or relief from symptoms, to help shift your focus away from your concerns. Remember that your healthcare provider is there to help you and ensure the best possible outcome. If you have specific concerns or questions about the ERCP procedure, don't hesitate to ask them for guidance and support. | 4 |
| I am worried about complications of ERCP, I am so stressed out and I don't know cope with all these. | I'm sorry to hear that you're feeling stressed and worried about the potential complications of ERCP. It's completely understandable to feel anxious about medical procedures, especially when there are risks involved. One way to cope with this anxiety is to make sure you have accurate and reliable information about the procedure and its potential complications. You can talk to your doctor or medical provider to get a clear understanding of what to expect during the procedure and what the risks and benefits are. It may also be helpful to talk to other people who have undergone ERCP and hear about their experiences. Online support groups or forums can be a good resource for this. In addition to gathering information, it's important to take care of yourself in the days leading up to the procedure. Make sure you're getting enough rest, eating well, and engaging in activities that help you feel relaxed and calm. You may also find it helpful to practice relaxation techniques like deep breathing or meditation. Remember that it's normal to feel anxious about medical procedures, but that doesn't mean you have to go through it alone. Don't hesitate to reach out to friends, family, or a mental health professional for support during this time. | 4 | I'm not a doctor, but I can try to provide you with some general information about ERCP (Endoscopic Retrograde Cholangiopancreatography) and ways to cope with stress related to medical procedures. Keep in mind that you should consult with a medical professional for advice tailored to your specific situation. ERCP is a procedure used to diagnose and treat problems in the liver, gallbladder, bile ducts, and pancreas. While it is generally considered safe, like any medical procedure, there are potential complications. Some possible complications of ERCP include: Infection Bleeding Pancreatitis Perforation of the intestine or bile ducts Adverse reaction to anesthesia or medications It's important to remember that these complications are relatively rare, and the procedure is performed by experienced medical professionals who take all necessary precautions to minimize risks. Here are some tips to cope with stress and anxiety related to medical procedures: Educate yourself: Understanding the procedure, its benefits, and potential risks can help you feel more in control and reduce anxiety. Communicate with your healthcare team: Share your concerns and ask questions. They can provide reassurance, clarify any misconceptions, and help you understand what to expect. Practice relaxation techniques: Deep breathing, progressive muscle relaxation, and meditation can help reduce stress and anxiety. Seek support: Talk to friends, family, or a mental health professional about your concerns. They can offer emotional support and help you put your fears into perspective. Focus on what you can control: Maintain a healthy lifestyle, follow your healthcare team's instructions, and engage in activities that promote relaxation and well-being. Remember, it's always best to consult with your healthcare provider if you have concerns or questions about an upcoming medical procedure. They can offer personalized advice and support tailored to your needs. | 5 |
| If I feel overwhelmed during this process of ERCP, I am anxious, sad, and confused! | It's completely understandable to feel overwhelmed, anxious, sad, and confused during a medical procedure like ERCP. It's important to remember that you're not alone, and there are steps you can take to cope with your feelings and make the process more manageable. Here are some tips that might help: Talk to your healthcare provider: Let your healthcare provider know how you're feeling. They may be able to offer you some reassurance and provide you with more information about the procedure. They may also be able to prescribe medication to help ease your anxiety. Practice relaxation techniques: Deep breathing, visualization, and meditation can help you feel calmer and more centered. You can try these techniques before and during the procedure. Bring a support person: Having someone you trust with you during the procedure can be comforting. This could be a family member or friend. Educate yourself: Sometimes, anxiety comes from not knowing what to expect. Ask your healthcare provider for information about the procedure, and do your own research if you feel comfortable doing so. Take care of yourself: Make sure you're eating well, getting enough sleep, and exercising regularly. Taking care of your physical health can help you feel more resilient and better able to cope with stress. Remember, it's normal to feel anxious and overwhelmed during medical procedures, but there are things you can do to help yourself feel better. Don't hesitate to reach out to your healthcare provider or loved ones for support. | 4 | It's normal to feel overwhelmed, anxious, sad, and confused during a medical procedure like an ERCP (Endoscopic Retrograde Cholangiopancreatography). To help you manage these emotions, consider trying the following strategies: Educate yourself: Learn as much as you can about the ERCP procedure, its purpose, and the potential risks and benefits. This will help you feel more informed and prepared. Talk to your healthcare team: Share your feelings and concerns with your healthcare team, including your doctor and the nurses involved in your care. They can provide information and reassurance to help ease your anxiety. Bring a support person: If possible, have a family member or friend accompany you during the procedure. They can provide emotional support and help you feel more comfortable. Practice relaxation techniques: Deep breathing exercises, mindfulness, and meditation can help you stay calm and focused during the procedure. Focus on positive thoughts: Remind yourself of the reasons you're undergoing the ERCP and the benefits it can provide. Focusing on the positive aspects of the procedure may help reduce negative emotions. Seek professional help: If your feelings of anxiety, sadness, or confusion become too much to handle, consider talking to a mental health professional. They can help you develop coping strategies and address your concerns. Remember that it's completely normal to feel a mix of emotions when facing a medical procedure. Communicating openly with your healthcare team and utilizing coping strategies can help make the experience more manageable. | 4 |
| Does my undergoing ERCP mean I have cancer? I am really worried about this! | No, undergoing an ERCP (Endoscopic Retrograde Cholangiopancreatography) does not necessarily mean that you have cancer. ERCP is a medical procedure used to diagnose and treat problems in the bile ducts and pancreas, such as gallstones, pancreatitis, or bile duct obstructions. During the procedure, a flexible tube with a camera on the end is inserted through the mouth and into the small intestine, allowing the doctor to examine the bile ducts and pancreas. If any abnormalities are found, the doctor may take a sample of tissue or fluid (biopsy) for further analysis. While cancer is one possible cause of abnormalities detected during an ERCP, it is not the only cause. Other conditions, such as infections or inflammation, can also be detected and treated through this procedure. It is important to discuss your concerns with your doctor, who can provide more information about the reasons for your ERCP and the possible outcomes of the procedure. | 3 | I understand that undergoing medical procedures can be concerning, but I am not a doctor and cannot provide medical advice or a diagnosis. An Endoscopic Retrograde Cholangiopancreatography (ERCP) is a procedure used to examine and treat conditions of the bile ducts, pancreas, and surrounding areas. It is not specifically a cancer diagnostic procedure, but can be used to identify and address various issues, such as gallstones, inflammation, or blockages. Having an ERCP does not necessarily mean you have cancer. Your doctor may have recommended the procedure for a variety of reasons. The best course of action is to discuss your concerns with your healthcare provider, as they will be able to give you the most accurate information about your specific situation. | 3 |

Endoscopy

<https://giendo.net/for-patients/endoscopy-faqs/>

<https://gi.org/topics/upper-gi-endoscopy-egd/>

<https://charlestongi.com/for-patients/faq/endoscopy-faqs/>

<https://www.insitedigestive.com/2021/07/09/endoscopy-procedure-faqs/>

<http://www.endoscopygastro.com/frequently-asked-questions/>

<https://digestivehealth.net/faqs/upperendoscopyfaqs.html>

Colonoscopy

<https://gi.org/patients/gi-health-and-disease/colonoscopy-faqs/>

<https://www.uofmhealth.org/conditions-treatments/digestive-and-liver-health/screening-colonoscopy/colonoscopy-faq>

<https://portlandgastro.com/colonoscopy-faqs-or-frequently-asked-questions/>

<https://www.beaumont.org/services/oncology/colorectal-cancer/colonoscopy-faqs>

<https://www.sutterhealth.org/services/gastroenterology/colonoscopy-faq-pamf>

<https://health.uconn.edu/gastroenterology/patient-care/patient-resources/colonoscopy-frequently-asked-questions/>

EUS

<https://www.asge.org/home/for-patients/patient-information/understanding-eus>

<https://my.clevelandclinic.org/health/diagnostics/12025-endoscopic-ultrasound>

<https://www.mayoclinic.org/tests-procedures/endoscopic-ultrasound/about/pac-20385171>

<https://patient.gastro.org/endoscopic-ultrasound-eus/>

<https://health.uconn.edu/gastroenterology/patient-care/patient-resources/colonoscopy-frequently-asked-questions/>

ERCP

<https://www.asge.org/home/for-patients/patient-information/understanding-ercp>

<https://www.niddk.nih.gov/health-information/diagnostic-tests/endoscopic-retrograde-cholangiopancreatography>

<https://www.hopkinsmedicine.org/health/treatment-tests-and-therapies/endoscopic-retrograde-cholangiopancreatography-ercp>

<https://www.mskcc.org/cancer-care/patient-education/patient-guide-endoscopic-retrograde-cholangiopancreatography-ercp>

<https://www.schneckmed.org/treatments/endoscopic-retrograde-cholangiopancreatography-ercp>

<https://health.uconn.edu/gastroenterology/patient-care/patient-resources/colonoscopy-frequently-asked-questions/>

**Methods**

**Data source**

We gathered frequently asked questions about endoscopic procedures from reputable professional societies and institutional web pages. (Supplemental file 1) Questions about endoscopic procedures, knowledge, and management were modified to ensure inclusiveness and representation of patients and caregivers. The criteria for exclusion consisted of questions that conveyed similar meanings, questions with ambiguous meanings (such as inquiries about how endoscopic procedures impact the body), queries that could differ from individual to individual (like the likelihood of a person's condition worsening after the procedure) and questions not related to the medical aspects of the procedures. A total of 117 questions were selected for common endoscopic procedures (EGD, colonoscopy, EUS, and ERCP). (Supplemental Table 1-4) Furthermore, we evaluated the capability of ChatGPT and GPT-4 to function as psychological support systems for patients (16 questions, Supplemental Table 5). Since there were no predefined benchmarks for the responses, the assessment of the model's effectiveness in responding to emotional support questions was performed by a board-certified psychiatric (LSM).

**Response generation**

Launched in November 2022, ChatGPT is a refined version of the GPT-3.5 Large Language Model, developed to advance Natural Language Processing. It was trained using a rich compilation of text data from diverse online sources up to 2021. Key to its design is incorporating user feedback, a strategy known as 'reinforcement learning from human feedback,' which allows it to generate more contextually apt and coherent responses. To mitigate abuse, ChatGPT developers have implemented multiple safeguards, such as preventing the generation of derogatory or harmful responses. Questions were input into the ChatGPT March 23 version. Each question was submitted as an independent separate prompt using the "New Chat" function. As in prior literature, every question was entered into ChatGPT twice independently by two authors, with both responses recorded to assess the reproducibility of ChatGPT's answers.. [8] We did not use GPT-4 for the primary analysis of this study as it is a paid subscription plan with a cap for the number of responses per hour and is not freely available to the public. [4]

**Grading of questions**

Review and grading of each response for EGD/Colonoscopy were conducted by at least two board-certified/eligible gastroenterologists (BPM and PP). Review and grading of each response for EUS/ERCP were conducted by at least two board-certified/eligible advanced endoscopists (IO and JE). The following grading system was employed to assess the precision of each response: 1. Comprehensive, 2. Correct but insufficient, 3. Mixed with correct and incorrect/outdated data, and 4. Completely incorrect, as done previously. [8] Each reviewer checked how similar the two answers were to each question to assess their reproducibility. If the responses differed, like having contradictory details or degrees of depth, reviewers separately evaluated both versions. Discrepancies in grading and assessment of reproducibility between the two reviewers were independently reviewed and resolved by a blinded third reviewer (KL).

**Emotional support questions and responses**

To address ChatGPT's performance on questions that addressed emotional support, two authors modified FAQs to address this need using questions posted on professional societies and institutional web pages (Supplemental Table 5). The emotional question was modified with a clear subject, relevant context, personalized touch, and open-endedness to elicit a response from ChatGPT. Performance of ChatGPT's potential as a psychological support system for patients was compared to the latest GPT-4 model by open AI. Since there was no established standard for the answers, a board-certified physician psychiatrist (LSM) participated in assessing the responses and gauging the model's effectiveness in providing emotional support. Questions were graded as follows: Not Comprehensive: The answer provided minimal information or needed to address the question adequately; Somewhat Comprehensive: The answer provided some information but left out important details; Moderately Comprehensive: The answer covered most aspects of the question but may have lacked depth or specificity in certain areas; Very Comprehensive: The answer addressed all aspects of the question and provided detailed information; Extremely Comprehensive: The answer was exhaustive, covering all aspects of the question with great detail and specificity.

**Statistical analysis**

A review was conducted to evaluate the reproducibility of ChatGPT's responses and compare the similarity of the two responses generated by the model for each question asked. Reproducibility was defined as the uniformity of two responses with matching grading categories. Both responses were graded when the responses for a specific question varied in content, such as having distinct cut-off values or meanings. The grading categories were divided into grades 1 and 2 vs. grades 3 and 4. If the two responses fell into separate groups, they were considered significantly different. For emotional support questions, the performance of ChatGPT was compared to GPT-4 by anonymizing the labels indicating which model is answering the questions. The proportions of each previously mentioned grade among responses for each domain of endoscopic procedures were computed and expressed as percentages (N%). The analyses were performed using STATA (version 16.1)
